# Latent Classes of Anthropometric Growth in Early Childhood Using Uni- and Multivariate approaches in a South African Birth Cohort

**DOI:** 10.1101/2023.09.01.23294932

**Authors:** Noëlle van Biljon, Marilyn T Lake, Liz Goddard, Maresa Botha, Heather J Zar, Francesca Little

## Abstract

**Background:** Conventional methods for modelling longitudinal growth data focus on the analysis of mean longitudinal trends or the identification of abnormal growth based on cross-sectional standardized z-scores. Latent Class Mixed Modelling (LCMM) considers the underlying heterogeneity in growth profiles and allows for the identification of groups of subjects that follow similar longitudinal trends.

**Methods:** LCMM was used to identify underlying latent profiles of growth for univariate responses of standardized height, standardized weight, standardized body mass index and standardized weight-for-length/height measurements and multivariate response of joint standardized height and standardized weight measurements from birth to five years for a sample of 1143 children from a South African birth cohort, the Drakenstein Child Health Study (DCHS). Allocations across latent growth classes were compared to better understand the differences and similarities across the classes identified given different composite measures of height and weight as input.

**Results:** Four classes of growth within standardized height (n_1_=516, n_2_=112, n_3_=187, n_4_=321) and standardized weight (n_1_=263, n_2_=150, n_3_=584, n_4_=142), three latent growth classes within Body Mass Index (BMI) (n_1_=481, n_2_=485, n_3_=149) and Weight for length/height (WFH) (n_1_=321, n_2_=710, n_3_=84) and five latent growth classes within the multivariate response of standardized height and standardized weight (n_1_=318, n_2_=205, n_3_=75, n_4_=296, n_5_=242) were identified, each with distinct trajectories over childhood. A strong association was found between various growth classes and abnormal growth features such as rapid weight gain, stunting, underweight and overweight.

**Conclusions:** With the identification of these classes, a better understanding of distinct childhood growth trajectories and their predictors may be gained, informing interventions to promote optimal childhood growth.

**Key Messages:**

- Four latent classes of growth were identified within standardized height and standardized weight.
- Three latent classes of growth were identified within standardized body mass index and standardized weight-for-length/height.
- Five latent classes of growth were identified within a multivariate response of standardized height and standardized weight.
- Latent classes identified using various composite measures of standardized height and standardized weight (standardized body mass index and standardized weight-for-length/height and a multivariate response of standardized height and standardized weight) were distinct, reiterating the benefit of examining each outcome.
- A strong association was found between various growth classes and abnormal growth features such as rapid weight gain, stunting, underweight and overweight.

## Introduction

A double burden of childhood malnutrition is emerging in low- and middle-income countries (LMICs) – undernutrition and obesity. According to UNICEF, in 2020, up to 23% of Southern African children under five years suffered from stunting, approximately 3% were wasted and 12% were overweight (1). Globally, more than two billion adults are obese, with over 70 percent of them residing in LMICs (2). Despite this high burden of malnutrition, data on early growth trajectories (defined as the first five years of life) are limited in LMICs.

Low birth weight (2500 grams or less), which is prevalent in LMICs, is associated with an increased risk of negative physical, cognitive and emotional health consequences throughout childhood (3). Given a low birth weight, the impact of catch-up growth (or rapid weight gain) in the short and long term is under debate (4). Considering short-term consequences, Victora et al., found catch-up growth to be associated with a reduced risk of hospitalisation (5). Meanwhile, children who experienced rapid weight gain between birth and 2 years have been found to be fatter with an increasingly central fat distribution at 5 years (6).

Specific growth patterns during infancy and childhood do not only have short-term consequences, but have also been shown to be associated with disease in later life, such as obesity (7), asthma, coronary heart disease (8), or stroke (9). Obesity, during childhood increases the risk of cardiovascular, metabolic and central nervous system disorders in later life (10). Stunting, an indicator of chronic malnutrition (11,12) has been shown to be associated with infections in childhood and obesity in adulthood, possibly due to a long-term impact on metabolic factors (13,14). Thus, longitudinal exploration of growth patterns during childhood is key to identifying individuals at greater risk of morbidity. The aim of this study was to identify heterogeneity in growth profiles in children from birth to five years to identify abnormal growth profiles of obesity, stunting, wasting or rapid weight gain in a South African birth cohort, the Drakenstein Child Health Study (DCHS).

To identify such distinct growth patterns, and subsequently children at greater risk of disorders in later life, approaches such as Growth Mixture Modelling (GMM), Latent Class Growth Analysis (LCGM), Latent Class Mixed Modelling (LCMM) and Time series clustering (TSC) (15–18) may be used. These aim to identify data-driven classes that share similar growth patterns. This paper refers to an LCMM approach to identify latent growth profiles for growth measurements from the Drakenstein Child Health Study.

The use of multivariate LCMM to identify latent growth classes based on joint height and weight measurements is proposed as this approach will not impose a restrictive, predefined relationship between Height and Weight as the use of zBMI or zWFH would when identifying distinct growth trajectories. No publications have identified latent classes using a multivariate response of Height and Weight or the comparison of latent growth class allocation across various growth responses. Additionally, an aim of this study was to assess the relationships across latent trajectories based on different growth responses through comparisons of subject allocation to the different groups, thus determining what additional information latent growth trajectories based on zBMI, zWFH or zHeight+zWeight may provide to those identified based on zWeight or zHeight alone. An expanded understanding of growth trajectories may identify vulnerable periods for intervention, to optimise child growth.

## Methods

The Drakenstein Child Health Study (DCHS) is a prospective South African birth cohort in which pregnant women were enrolled between March 2012 and March 2015 and followed through birth and childhood. The DCHS aims to investigate the early life determinants of child health and mechanisms underlying the development of disease, including environmental, nutritional, infectious, psychosocial, and maternal physical and mental health determinants. These measures have been longitudinally collected from the antenatal period through childhood (19). DCHS has detailed comprehensive growth measurements from birth through childhood in a LMIC setting. Measurements of Height, Weight, Mean-Upper Arm circumference, Triceps Skin Fold Thickness, Head Circumference, Body Mass Index and Weight-for Height have been recorded.

This paper focuses on a model-based approach to identify latent growth profiles for standardized height and weight measurements, as well as the composite measures of these responses, namely standardised Body Mass Index (zBMI), standardised Weight-for-Height (zWFH) and a multivariate response of zHeight and zWeight (zHeight+zWeight).

### Participants

The DCHS enrolled women, from a poor peri-urban area in South Africa, during their second trimester (20-28 weeks) of pregnancy. Mother-child pairs were followed for up to at least 10 years of child age. For this paper, growth measures from birth until 5 years were included. Children were followed and measurements performed at birth and at 6 weeks, 10 weeks, 14 weeks, 6 months, 9 months, and 12 months then 6 monthly until 5 years (Figure 1; Figure A1).

**Figure 1:**
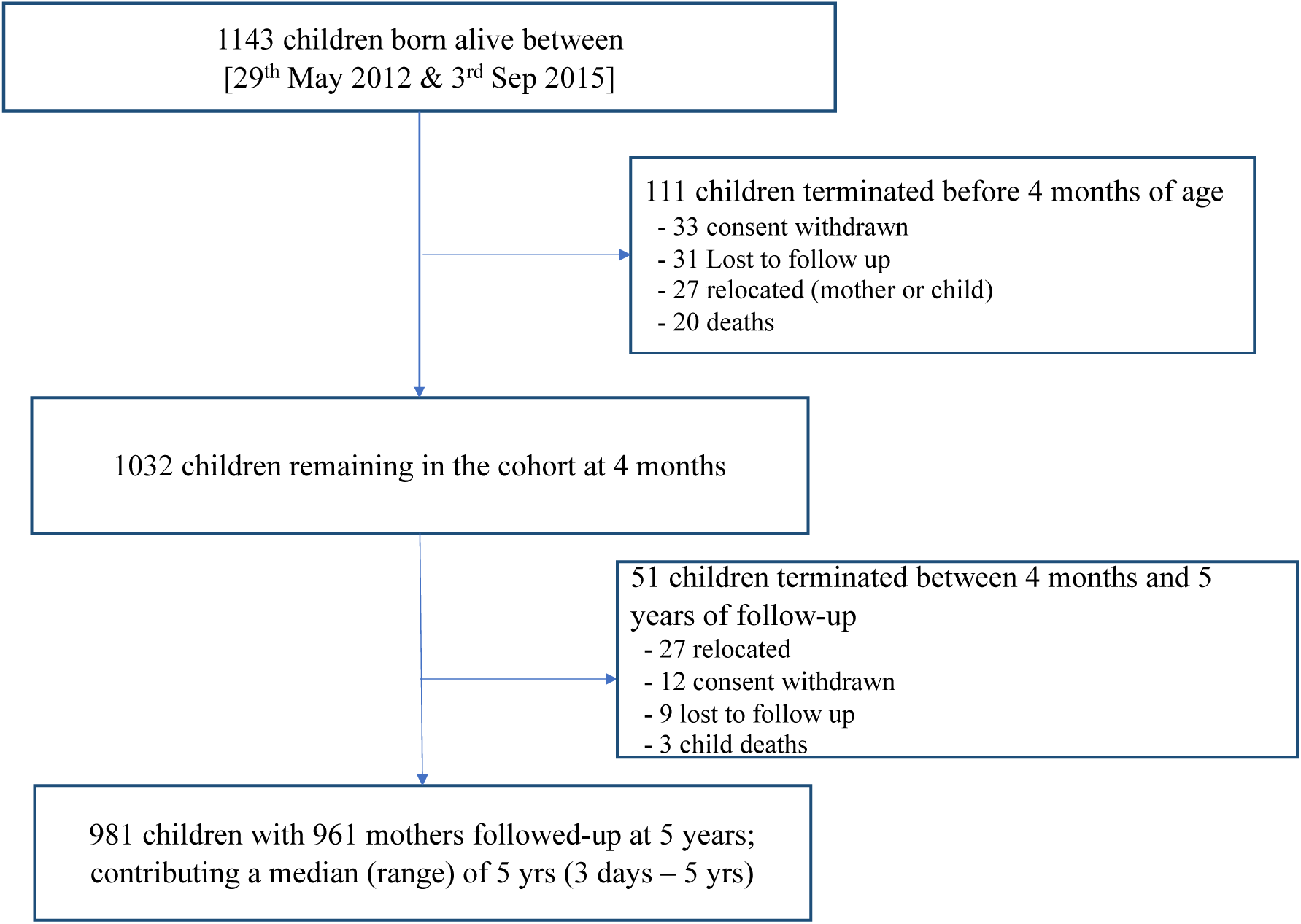
The DCHS Cohort of children from birth until 5 years of age.

### Outcomes

Growth measurements included Height and Weight from which standardised Body Mass Index (zBMI) and standardised Weight for Height (zWFH) were calculated. Height (recumbent length (recorded in centimetres) measured as distance from crown to foot using a Seca length-measuring mat (Seca; Hamburg, Germany) from birth until 24 months, after which this was recorded as standing height using a wall-mounted stadiometer (Panamed; Philippines) and weight (the mass of a subject (to the nearest 10 g) in light or no clothing using a Tanita digital platform scale (TAN1584; IL, USA)) were measured by trained study staff. Each measurement was taken twice per visit per child to serve as technical replicates. Measures within 0.5cm and 0.1kgs of each other for height and weight respectively were acceptable and the first measurement was used. If measures were not within this acceptable range, a third measurement was taken and used. For comparability, standardised growth responses (with respect to Growth References) were analysed within this report; these are denoted using “zGrowthResponse” such as zHeight for standardised height and zWeight for standardised weight. Growth responses as well as the number of observations per visit are summarised in Table A1.

Fenton growth references were used to calculate standardised growth scores for prematurely born children (before 37 weeks’ gestation); for full term infants WHO references adjusted for gestational age were used until two years (as is the convention), (20–22). Standardised height (zHeight; derived using WHO and Fenton reference ranges), standardised weight (zWeight; derived using WHO and Fenton reference ranges), standardised body mass index (zBMI; weight in kg divided by height in meters squared, standardised using WHO reference ranges) and standardised Weight for Length/ Height (zWFH; derived using WHO reference ranges) were calculated.

To identify children that experienced rapid weight gain, a change in zWeight equal or greater than 0.67 within the first nine months of life was used, as per convention, although there is disagreement on the optimal time frame to use (23).

### Statistical Analysis

The modelling of the respective growth measures involved two overarching approaches: 1) Univariate- and 2) Multivariate Latent Class Mixed Models. Within the univariate modelling section each growth measure was described and analysed independently. The responses considered with this approach were zHeight, zWeight, zBMI and zWFH. The multivariate approach allowed the growth measures to be analysed together, thus identifying groups of subjects that follow similar trajectories while considering more than one growth response. Using this approach, latent classes from a model based on zHeight and zWeight was considered as an alternative to latent classes from the individual models for the calculated composite scores, zBMI and zWFH.

Prior to LCMM, univariate mixed effect models for the mean response were fit to each individual growth measure to determine the optimal structure for modelling the association with time. Both linear and non-linear relationships between time and the growth measurements were considered to ensure the best model fit, including linear, cubic splines, fractional polynomials, and piecewise linear splines. For growth measurement these four different model formulations with differing knot choices for the spline options were compared to each other. The piece-wise linear spline formulation was chosen for inclusion in the latent trajectory modelling based on model fit and ease of interpretation. Thus, for each growth measurement, a piece-wise linear spline was used to describe the change in the measure with increasing age. The different model formulations and the location and number of knots were chosen to minimise AIC and thus allow for the best fit possible. Details of the final form of the mixed effect model for each of the responses are found within the supplementary materials – Part 2: Extended Statistical Methods.

Latent growth classes within zHeight, zWeight, zBMI and zWFH were identified using the latent class mixed model (LCMM) approach (18). LCMM makes use of three models: a structural model, taking the form of a linear mixed effect model which describes the association with time, a measurement model, which relates the latent process to the observations and a multinomial logistic model, which describes class membership. This is described in greater detail within the supplementary materials. Latent classes were identified without adjusting for any covariates. The choice of the number of latent classes was accomplished through the use of various fit statistics. For each growth measure, the latent class mixed effect model was fit with class numbers ranging from 1 to 6. The number of classes resulting in the lowest BIC (a measure of model fit to the data), largest Entropy (a measure of the level of discrimination between the classes), and lowest Integrated Classification Likelihood (ICL, a measure of fit conditional on class discrimination) were selected subject to their size being greater than 5% of the sample and their stability.

The stability of the identified classes and profiles thereof were validated through internal cross-validation. This validation step involved randomly selecting approximately 50% of the subjects and refitting the latent class mixed effect model with the chosen number of classes. This was repeated with 10 different random samples. The response profile of the classes identified were plotted together on one set of axes and the class number that appeared to have the most consistency with respect to the class trajectories was taken into consideration in addition to the fit statistics.

LCMM can be extended to model multiple responses simultaneously by fitting multivariate mixed models and multivariate latent class mixed models for multivariate longitudinal response variables. This approach was used to identify latent growth classes within the combined responses of zHeight and zWeight, defining each longitudinal dimension as a latent process (Figure 2), to allow for more flexible relationships between zHeight and zWeight within the multivariate setting (18). The model formulation included two structural models as well as two measurement models, each with response specific parameter estimates, with one multinomial logistic model still determining the probability of allocation to the respective classes (Figure 2). The likelihood is a function of the individual structural and measurement models as well as the multinomial logistic class allocation model and hence maximisation of the likelihood yields class allocations that are dependent on all three components. For this multivariate response model, linear transformations were used as link functions within the measurement model, though various transformations were considered that produced similar results and hence the simpler linear link was chosen.

**Figure 2:**
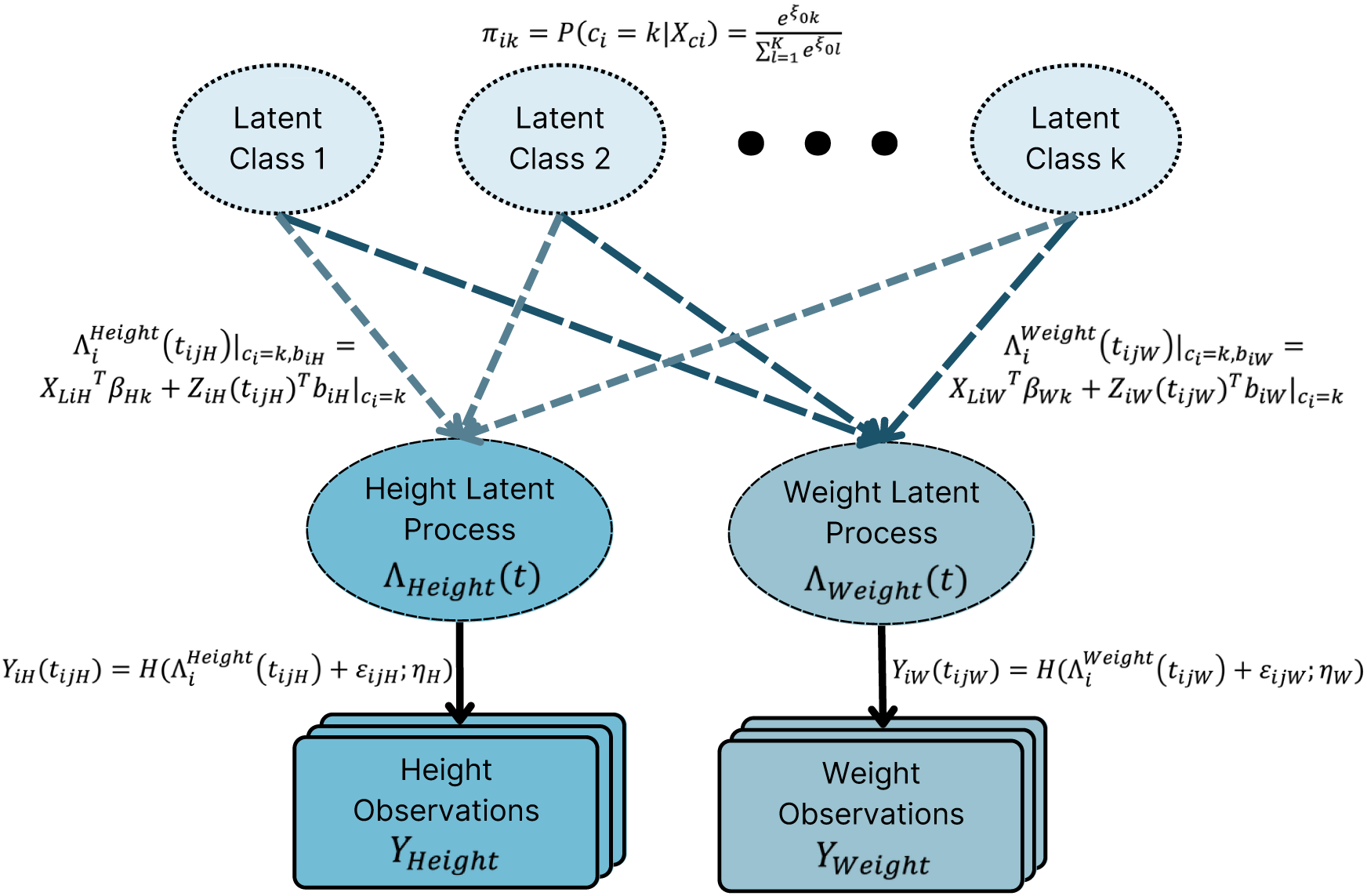
Diagram illustrating the structure of the multivariate LCMM process when considering distinct latent structures for zHeight and zWeight responses respectively.

Additional detail as well as specification of terms can be found within the Supplementary Materials – Part 2.

## Results

Amongst 1137 mothers enrolled there were 1143 children live born, with 981 (85.8%) followed through 5 years of age with detailed measurements as shown in Figure 1. Twenty-three (2%) children died in infancy and 139 (12.2%) children were lost to follow up, the majority of which occurred before 6 months of age. All growth measurements were recorded at each visit, however some measurements were excluded due to instrumental or measurement error.

### Study Cohort

Six-hundred and ninety-five (60.8%) of the mothers did not complete secondary schooling education, while 988 (86.4%) were members of households where the average monthly income was below R5000 (approximately USD276). One hundred and ninety-one (16.7%) children were born prematurely, the majority late preterm (born between 34-37 weeks gestational age). There were 247 (21.6%) HIV-exposed but uninfected children, with only 2 HIV-infected. Characteristics of included children were similar to those excluded; Table A2 summarises the total, included and excluded cohort with respect to maternal, child and socio-economic characteristics.

### Average Trajectory of Growth Responses

Figure 3 illustrates the standardised growth responses from birth until five years. As these observations represent standardized growth scores, the average trajectory should follow the line of y=0. On average, lower zHeight and zWeight scores from what were anticipated according to WHO references were observed. Larger than expected zBMI and zWFH scores were observed, indicating that while children were below expected weight, their height deviated even further below expectation, resulting in above expected zBMI and zWFH. The trajectories of the unstandardised, observed, growth responses are illustrated within Figure A2 and Table A1 in the supplementary materials.

**Figure 3:**
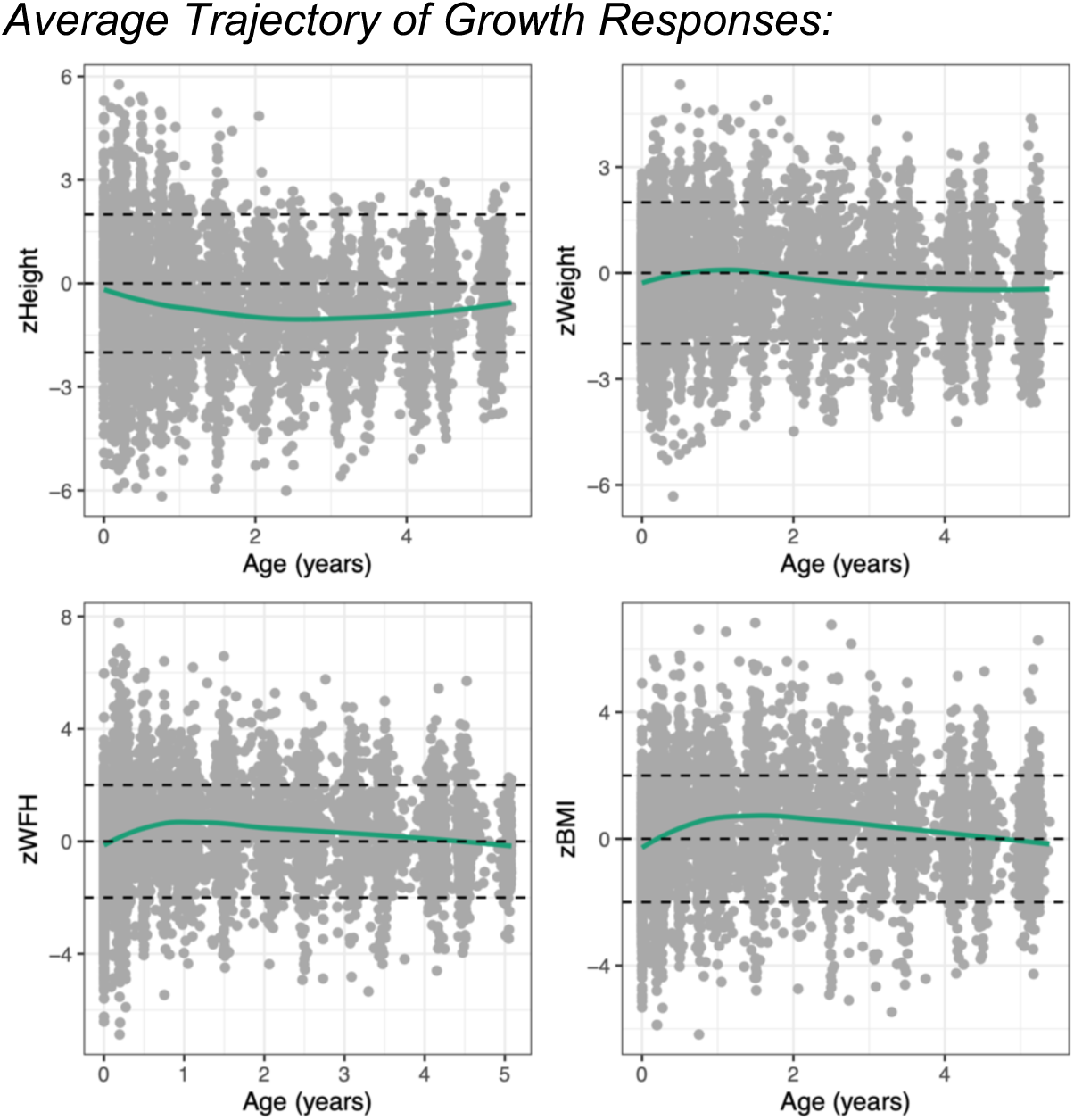
Standardised growth measures from Birth until age 5 with a smoothed average trajectory.

### Describing Growth Responses over Time

The zHeight and zWeight, zBMI and zWFH standardized measurements over time were modelled using mixed effect models with piecewise linear splines to capture the time component. Between 3 and 4 knots were placed between ages 0.25 and 2.5 years for the different growth measures and the resulting mean profiles are illustrated in the supplementary materials (Table A3; Figure A3). The early placement of the knots with no knots beyond age 2.5 reflects that most of the changes in trajectory structure occurred before two years of age. These changes in trajectory structure may also be a feature of more frequent data collection during the infancy. To determine whether the choice of knot location impacted the subsequently identified latent profiles, the longitudinal profiles given additional knots placed at later time points (ages 0.25,0.75,1,1.5,2,3,4; Figure A4) were compared to those described in Figure A3. The profiles did not show clear deviation, and thus the knot locations specified in Table A3 were used.

### Selecting K, the number of latent classes

With the previously described broken stick specifications of the longitudinal process, latent class mixed models were fit to each of the respective growth measures, zHeight, zWeight, zBMI and zWFH, individually. Additionally, this approach was also applied to the multivariate response, zHeight + zWeight. An important step within this process was choosing the appropriate number of latent classes, k.

Figure 4 illustrates the process whereby the number of latent classes (k) for the trajectories of zHeight was selected. Figure 4A shows the fit statistics for 1 to 5 latent classes. In this case the more classes, the lower the BIC. Similarly, the entropy for k=5 was the greatest (Figure 4A). The class sizes when five latent classes were considered were still greater than 5%, however the Figure 4D shows a lack of stability when k=5. As a compromise, k=4 was chosen as the optimal class number as the fit statistics were very similar to that of k=5, while the stability shown in Figure 4C is consistent.

**Figure 4:**
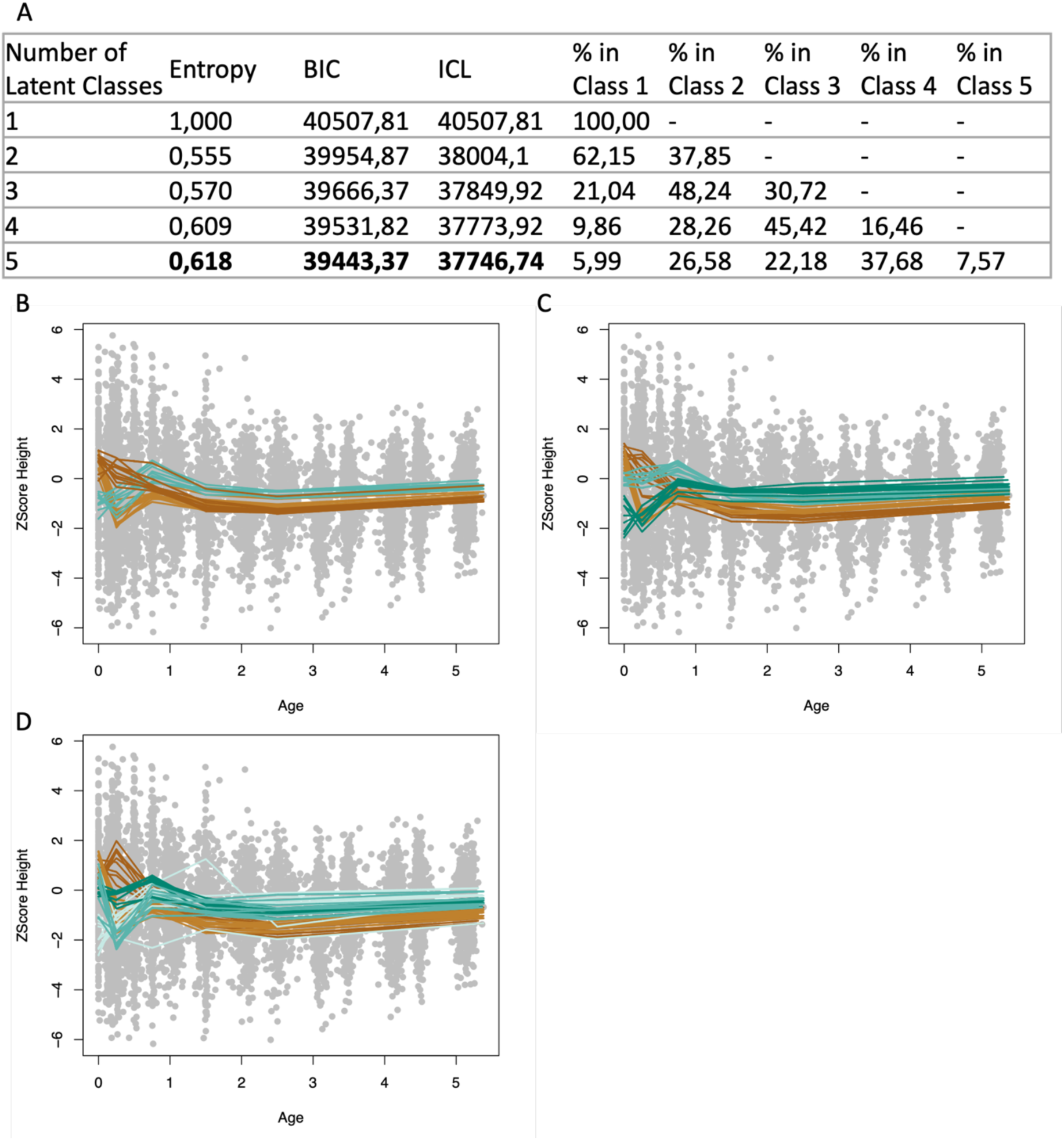
Information used to choose the appropriate value for k, the number of latent classes within standardised Height. A) Fit statistics for k=(1:5). Profiles of LCMM Classes identified within standardised Height using randomly selected 50% of subjects, repeated 10 times for B) k=3, C) k=4 and D) k=5.

This process was repeated for all growth responses, Figures A5-A9 in the supplementary materials. For zWeight, four latent classes were identified as optimal (Figure A5). Three latent classes were identified as optimal within zBMI and zWFH (Figure A6, A7). When considering the multivariate zHeight+zWeight model, five latent classes were identified as optimal (Figure A8-A9).

### Growth trajectories: Univariate trajectories

Each response, described using the broken stick model previously indicated, fit with the latent class mixed model given the optimal number of k is illustrated in Figure 5, thus illustrating the trajectories of each latent growth class identified within the respective growth responses. The model estimates from the fixed effect components can be found within Tables A4-A9 in the Supplementary Materials.

**Figure 5:**
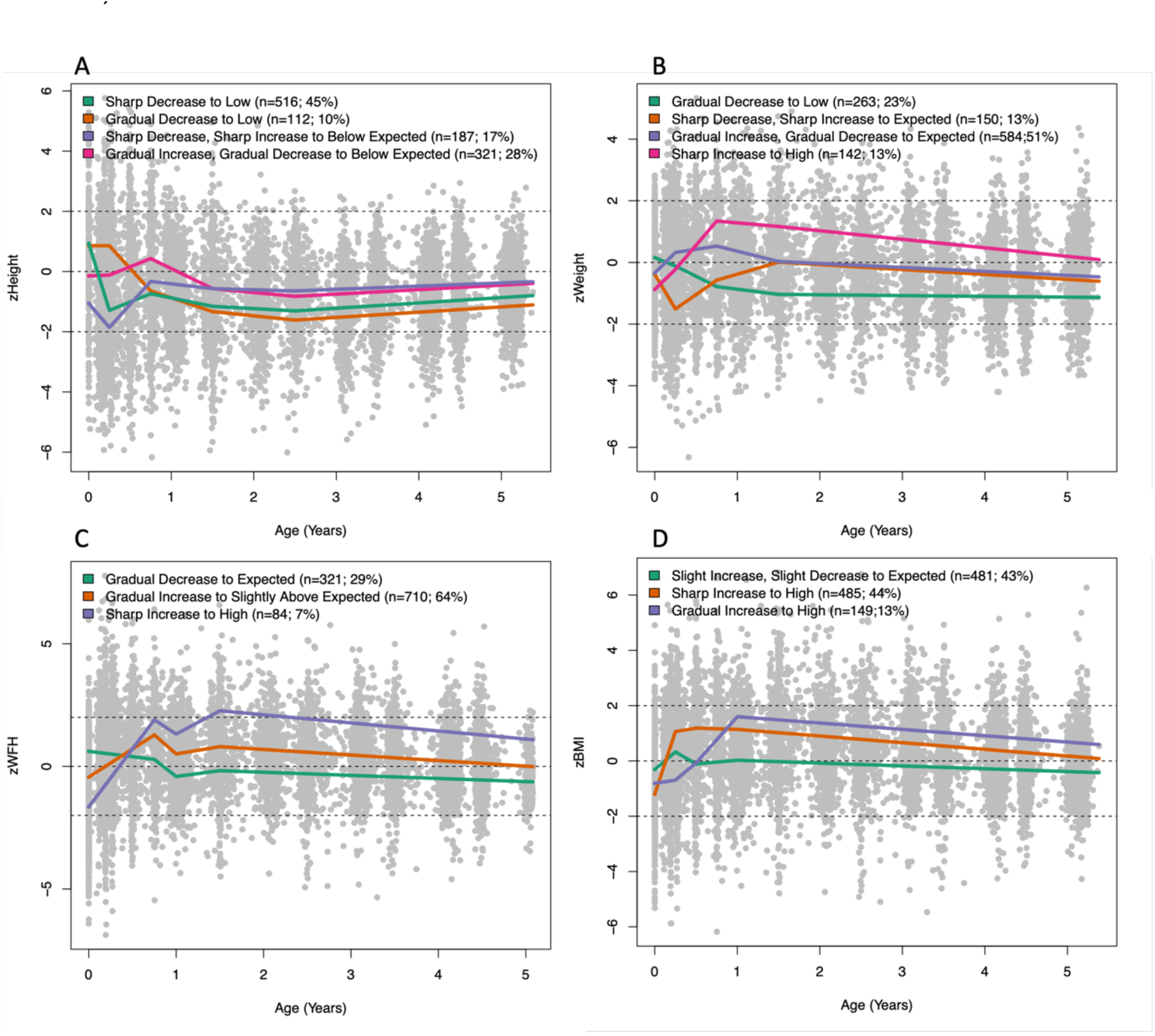
The average trajectories of latent classes identified within A) zHeight, B) zWeight, C) zWFH and D) zBMI from birth until age of five.

### zHeight

The four latent classes for zHeight are illustrated in Figure 5(A). The following features were observed: Firstly, zHeight profiles below the expected are observed (this was also reflected in zWeight profiles). Secondly, much of the differences in structure of trajectories occur before 1.5 years, after which the four trajectories settle at different levels. The same result was observed when an increased number of knots were placed at locations after 1.5 years; these results are shown within the supplementary materials Figure A10-A15. This change in profiles prior to and after 1.5 years is reflected in the early changes in slope and later levels naming of the four identified classes (for example, sharp increase to expected levels). Two classes that settled slightly closer to the expected height are seen, one class of 321 (labelled “Gradual Increase, Gradual Decrease to Below Expected”) children who started at expected height, increased to slightly higher heights than expected and then settled at a level of less than one standard deviation below the expected level. Another class (labelled “Sharp Increase, Sharp Decrease to Below Expected”) includes 187 children who started with lower heights than expected but by the age of 1 year have caught up to a level less than one standard deviation below the expected height. A further two classes show children who were taller than expected at birth, but who then attained a level up to two standard deviations below the expected height, one group of 516 children (labelled “Sharp Decrease to Low”) earlier than the other group of 112 children (labelled “Gradual Decrease to Low”).

### zWeight

Four latent classes were identified within zWeight. Similar to zHeight, the heterogeneity in structure is seen in the profiles prior to 1.5 years. Two classes (labelled “Gradual Increase, Gradual Decrease to Expected” and “Sharp Decrease, Sharp Increase to Expected”) initialize close to the expected zWeight at birth and diverge within the first 1.5 years, one class slightly above expected zWeight and another dipping well below expected, after which they meet and show subjects that follow expected weight for age from 1.5 to 5 years old (Figure 5(B)). Two classes with birthweights as expected who then deviated from expected weight profiles were observed, resulting in a class with above expected weights (labelled “Sharp Increase to High”) and a class with lower-than-expected weights (labelled “Gradual Decrease to Low”).

### zWFH

Three classes were identified for zWFH (Figure 5(C)). A class that almost follows the expected weight-for-height from birth until five years (labelled “Gradual Decrease to Expected”), and two classes with consistently greater weights given their height and sex. The group with the greatest weight for height (labelled “Sharp Increase to High”), showed the smallest weight-for-height at birth.

### zBMI

Three classes were identified for zBMI, all of which were characterised by early increasing zBMI measurements (Figure 5(D)). A class that closely follows the expected zBMI from birth until five years, except for a brief fluctuation above the expected BMI before 6 months of age (labelled “Sharp Increase, Sharp Decrease to Expected”). Two classes with smaller than expected zBMI at birth which both settle above the expected zBMI. One class settles earlier on, at 3 months of age (labelled “Sharp Increase to High”) while the second settles around one year of age (labelled “Gradual Increase to High”).

Figure 5 interestingly also show that those with the most extreme lower than expected measurements at or soon after birth, settled at the highest levels, and vice versa, indicating an over-correction of sorts.

### Multivariate trajectories

Figure 6 illustrates the zHeight and zWeight profiles for the five classes determined for the joint zHeight+zWeight measurements. Three hundred and eighteen children were allocated to a class with low zHeight and zWeight from birth until five years (labelled “i”). Both zHeight and zWeight was above expected at birth but slowly decreased to below expected at birth for 205 children (labelled “ii”). A small group of children (n=75) showed a delay in both zHeight and zWeight in the first few months of growth, however this did not appear to have long lasting impacts on zHeight or zWeight trajectories (labelled “iii”). A class of 296 children illustrate weight and height measurements equal to what was expected prior to age 1 however in subsequent observations these children were shorter than expected given their age and sex, while maintaining an expected weight (labelled “iv”). The last identified class shows individuals with a low zWeight at birth which steadily increased within their first year and remained above what was expected (labelled “v”). This same group was also shorter than expected at birth, with a slight delay in height growth during the first year of life, after which they followed expected height for age and sex.

**Figure 6:**
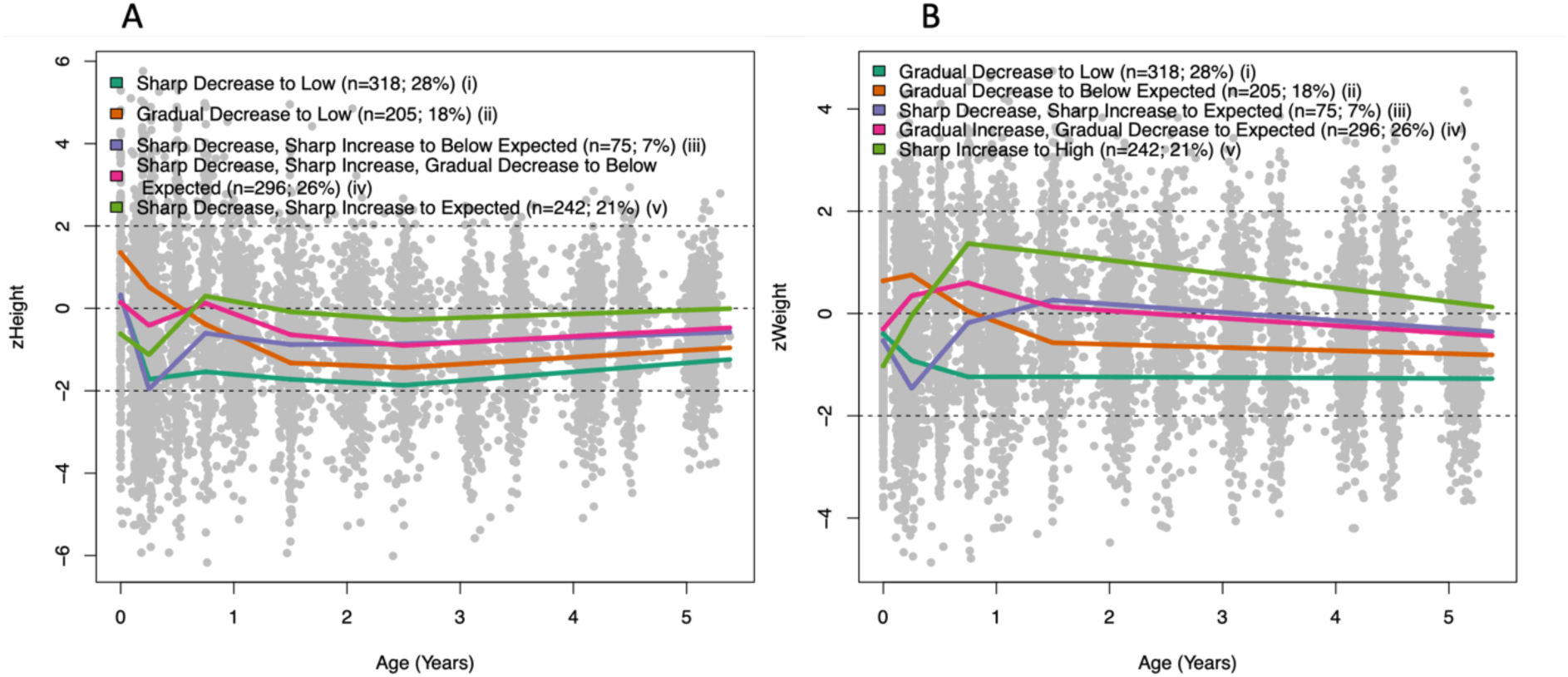
The average trajectories of four classes identified within zHeight+zWeight from birth until age of five, illustrated using A) zHeight and B) z Weight.

The accordance or dissimilarity of zHeight and zWeight trajectories within the latent classes is of interest. Class (iv) shows different profiles in particular before 1.5 years where an increasing zWeight and decreasing zHeight can be seen. Additionally, class (v) also shows an increasing zWeight with decreasing zHeight within the first few months of life.

To determine whether the choice of knot location impacted the subsequently identified latent profiles, the latent class identification process was repeated with additional knots placed at later time points. The extended knot locations used were (0.25,0.75,1,1.5,2,3,4) years for all responses considered. The results from this longitudinal specification are presented in the supplementary materials (Figure A16-A17). The results did not differ, and thus the knot locations specified in Table A3 were used.

### Comparison of Latent Class Allocations

The correspondence between latent class allocations for individual children based on different growth measurements and on allocations based on univariate versus multivariate models are shown in Tables 1-3 and A10. Table 1 illustrates the correspondence between allocations based on zHeight, zWeight and zHeight and zWeight within a joint model in a three-way contingency table. In contrast, Tables 2-3 summarise the agreement between composite measures of height and weight (zBMI and zWFH) and the class allocations based on models for joint zWeight and zHeight. Finally, Table A10 summarises the agreement between the latent classes identified given zWFH or zBMI.

**Table 1:** Tabulation of multivariate group allocation versus.

(a) univariate height group allocation
| Multivariate | zHeight: | Gradual Decrease to Low | Gradual Increase; Gradual Decrease to Below Expected | Sharp Decrease to Low | Sharp Decrease; Sharp Increase to Below Expected | All Height |
| --- | --- | --- | --- | --- | --- | --- |
| i: zHeight sharp decrease to low<br>zWeight gradual decrease to low |  | 10(3.14%) | 48(15.09%) | 218(68.55%) | 42(13.21%) | 318(100%) |
| ii: zHeight gradual decrease to low<br>zWeight gradual decrease to below expected |  | 84(41.0%) | 46(22.4%) | 75(36.6%) | 0(0%) | 205(100%) |
| iii: zHeight sharp decrease, sharp increase to below expected<br>zWeight sharp decrease, sharp increase to expected |  | 3(4.0%) | 12(16.0%) | 42(56.0%) | 18(24.0%) | 75(100%) |
| iv: zHeight sharp decrease, sharp increase, gradual decrease to below expected,<br>zWeight gradual increase, gradual decrease to expected |  | 12(4.1%) | 138(46.6%) | 128(43.2%) | 18(6.1%) | 296(100%) |
| v: zHeight sharp decrease, sharp increase to expected,<br>zWeight sharp increase to high |  | 3(1.3%) | 77(31.8) | 53(21.9%) | 109(45%) | 242(100%) |
| TOTAL |  | 112(9.9%) | 321(28.3%) | 516(45.4%) | 187(16.5%) | 1136(100%) |

(b) univariate weight group allocation
| Multivariate | zWeight: | Gradual Decrease to Low | Gradual Increase; Gradual Decrease to Expected | Sharp Decrease; Sharp Increase to Expected | Sharp Increase to High | All Weight |
| --- | --- | --- | --- | --- | --- | --- |
| i: zHeight sharp decrease to low<br>zWeight gradual decrease to low |  | 138(43.4%) | 118(37.1%) | 61(19.2%) | 1(0.3%) | 318(100%) |
| ii: zHeight gradual decrease to low<br>zWeight gradual decrease to below expected |  | 105(51.2%) | 100(48.8%) | 0 | 0 | 205(100%) |
| iii: zHeight sharp decrease, sharp increase to below expected,<br>zWeight sharp decrease, sharp increase to expected |  | 2(2.7%) | 16(21.3%) | 51(68%) | 6(8%) | 75(100%) |
| iv: zHeight sharp decrease, sharp increase, gradual decrease to below expected,<br>zWeight gradual increase, gradual decrease to expected |  | 18(6.1%) | 265(89.5%) | 5(1.7%) | 8(2.7%) | 296(100%) |
| v: zHeight sharp decrease, sharp increase to expected,<br>zWeight sharp increase to high |  | 0 | 82(33.9%) | 33(13.6%) | 127(52.5%) | 242(100%) |
| TOTAL |  | 263(23.2%) | 581(51.1%) | 150(13.2%) | 142(12.5%) | 1136(100%) |

**Table 2:**
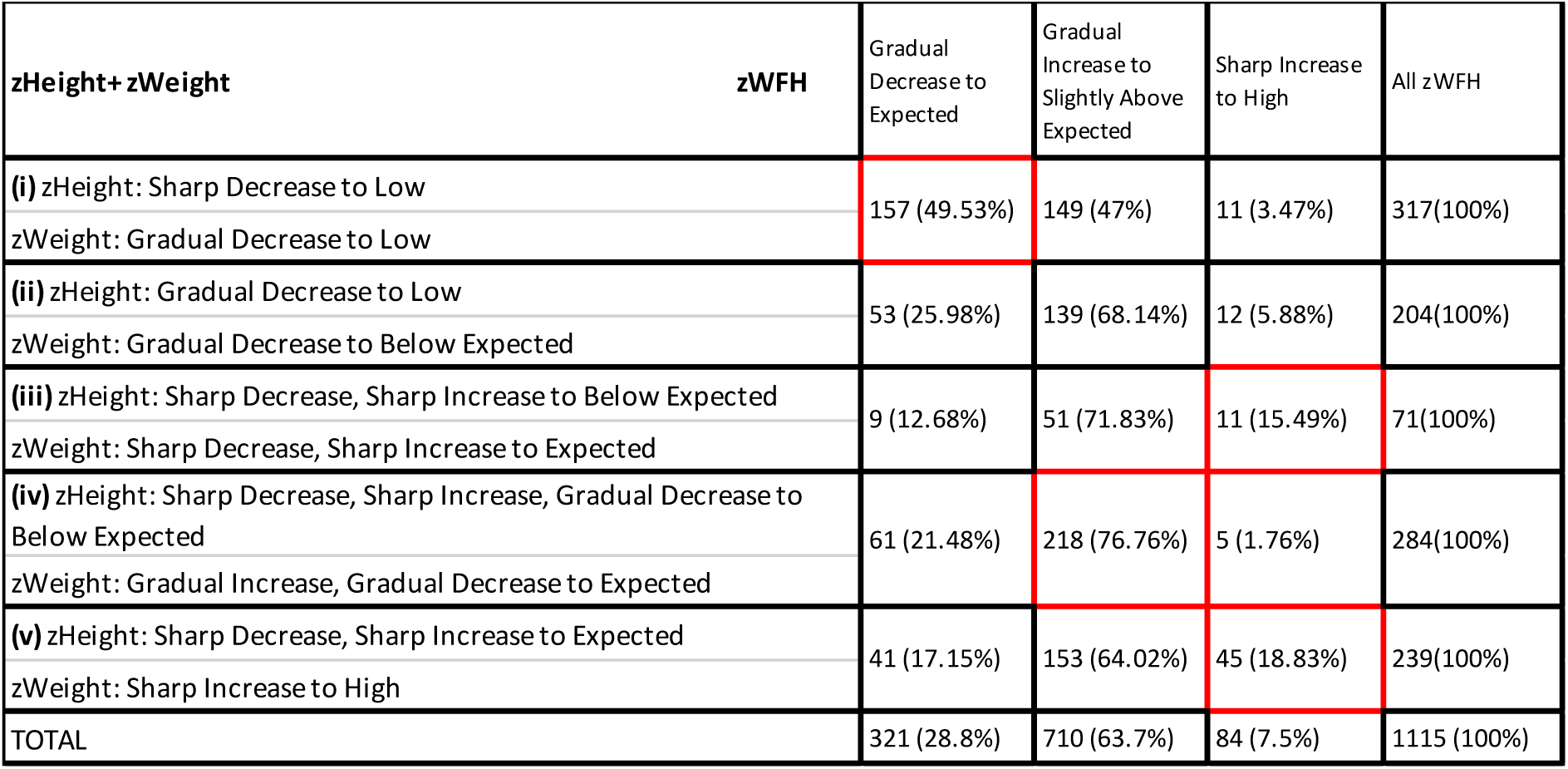
Comparison of subject allocations to zWFH and multivariate zHeight + zWeight classes. Comparing these allocations when considering trajectories from birth until 5 years of age.

**Table 3:** Comparison of subject allocations to zBMI and multivariate zHeight + zWeight classes. Comparing these allocations when considering trajectories from birth until 5 years of age.

| **zHeight+ zWeight** | **zBMI** | Gradual Increase to High | Sharp Increase to High | Slight Increase, Slight Decrease to Expected | All zBMI |
| --- | --- | --- | --- | --- | --- |
| **(i)** zHeight: Sharp Decrease to Low<br>zWeight: Gradual Decrease to Low |  | 29 (9.15%) | 89 (28.08%) | 199 (62.78%) | 317 (100%) |
| **(ii)** zHeight: Gradual Decrease to Low<br>zWeight: Gradual Decrease to Below Expected |  | 17 (8.33%) | 88 (43.14%) | 99 (48.53%) | 204 (100%) |
| **(iii)** zHeight: Sharp Decrease, Sharp Increase to Below Expected<br>zWeight: Sharp Decrease, Sharp Increase to Expected |  | 23 (32.39%) | 20 (28.17%) | 28 (39.44%) | 71 (100%) |
| **(iv)** zHeight: Sharp Decrease, Sharp Increase, Gradual Decrease to Below Expected<br>zWeight: Gradual Increase, Gradual Decrease to Expected |  | 17 (5.99%) | 155 (54.58%) | 112 (39.44%) | 284 (100%) |
| **(v)** zHeight: Sharp Decrease, Sharp Increase to Expected<br>zWeight: Sharp Increase to High |  | 63 (26.36%) | 133 (55.65%) | 43 (17.99%) | 239 (100%) |
| **TOTAL** |  | 149 (13%) | 485 (44%) | 481 (43%) | 1115 (100%) |

### Comparison of Univariate and Multivariate zHeight and zWeight latent class allocations

Tables 1a and b cross-tabulate the multivariate allocations to the univariate allocations of subjects based on Height and Weight, respectively. These tables show the number of children in each cross-tabulation and percentages of the multivariate classifications allocated to the different univariate height or weight classes. When cell frequencies exceed the distribution of the total cohort across the height or weight classes, they are indicative of a strong association with the specific multivariate class.

There is a strong agreement between identified trajectories such as within multivariate class (i) where the zHeight and zWeight classes with the strongest association describe the same trajectories as described by multivariate class (i):

Multivariate Class (i), Trajectories:

- zHeight shows a Sharp Decrease to Low,
- zWeight shows a Gradual Decrease to Low. Univariate Trajectories:

`o` zHeight shows a Sharp Decrease to Low,
`o` zWeight shows a Gradual Decrease to Low.

Multivariate classes (iii) and (v) showed identical zWeight profiles when comparing the univariate and multivariate classes with the strongest associations, while the zHeight trajectories for these classes differed between the multivariate and univariate allocations.

Multivariate Class (iii), Trajectories:

- zHeight shows a Sharp Decrease, Sharp Increase to Below Expected,
- zWeight shows a Sharp Decrease, Sharp Increase to Expected. Univariate Trajectories:

`o` zHeight shows a Sharp Decrease to Low,
`o` zWeight shows a Sharp Decrease; Sharp Increase to Expected.

Multivariate Class (v), Trajectories:

- zHeight shows a Sharp Decrease, Sharp Increase to Expected,
- zWeight shows a Sharp Increase to High. Univariate Trajectories:

`o` zHeight shows a Sharp Decrease; Sharp Increase to Below Expected,
`o` zWeight shows a Sharp Increase to High.

For multivariate class (ii), the converse of this was observed where the zHeight profiles descriptions based on multivariate and univariate allocations with the strongest association were identical, while the zWeight profiles differ with respect to the level at which the profiles settled.

Multivariate Class (ii), Trajectories:

- zHeight shows a Gradual Decrease to Low,
- zWeight shows a Gradual Decrease to Below Expected. Univariate Trajectories:

`o` zHeight shows a Gradual Decrease to Low,
`o` zWeight shows a Gradual Decrease to Low.

Finally, multivariate class (iv) showed identical zWeight profiles across univariate and multivariate allocations with the strongest association, while the zHeight profiles differ both with respect to level and rate of change prior to 1.5 years:

Multivariate Class (iv), Trajectories:

- zHeight shows a Sharp Decrease, Sharp Increase, Gradual Decrease to Below Expected,
- zWeight shows a Gradual Increase, Gradual Decrease to Expected. Univariate Trajectories:

`o` zHeight largely described by two classes, one shows a Gradual Increase; Gradual Decrease to Below Expected, the second shows a Sharp Decrease to Low,
`o` zWeight shows a Gradual Increase; Gradual Decrease to Expected.

In summary, it appears that based on the association across the univariate and multivariate class allocations, zWeight profiles appear to be in agreement more frequently than zHeight allocations. Additionally, the multivariate approach allowed for flexible profiles of one measurement (say zHeight) while holding the profiles for the other measurement (say zWeight) constant. For example, multivariate classes (i) and (ii) both reflect gradual decreases for zWeight but class(i) capture a sharp decrease in zHeight while class(ii) allowed for a more gradual decrease in zHeight. Class (ii) additionally also included a significant number of children whose weights first increased before showing a gradual decrease to their expected weight. None of these associations were perfect, making it difficult to get a clear assessment of the additional information conveyed by the multivariate approach.

When considering the link functions of this multivariate model that connects the observed process to the latent process, the slope of the zWeight link covers a greater range of the longitudinal process, thus the influence of weight may thus also be greater in predicting class allocations (Figure A19). A cubic function with three equally spaced knots was also considered as a more appropriate link function, while the fit characteristics produced were slightly better, the final profiles identified were almost identical (Figure A19) and thus the less complex linear link was used.

### Comparison of class allocations based on composite measures (zWFH and zBMI) to that based on joint zHeight and zWeight measurements

Sixty Four percent of children were allocated to the “Gradual Increase to Slightly Above Expected” zWFH class, while based on the joint zHeight + zWeight response a large proportion of these children were allocated to all five classes (Table 2); this indicates that the zHeight + zWeight approach is able to separate children into growth classes that would have otherwise been lost if only zWFH was used as input. Interestingly, 49.5 percent of individuals allocated to zHeight+zWeight class (i) were allocated to the zWFH “Gradual Decrease to Expected” class, which is substantially higher than the overall percentage, 29%, of individuals allocated to this class, thus showing a strong agreement between these profiles. However, looking at the descriptions of these trajectories, the multivariate profiles are described as a “Sharp Decrease to Low” within zHeight and “Gradual Decrease to Low” within zWeight while the zWFH trajectory does not suggest low zWFH. Thus, while an individual may be identified with abnormal growth through zWeight or zHeight, the use of zWFH may miss such a diagnosis.

In Table 3, the “Slight Increase, Slight Decrease to Expected” zBMI class shows a strong association with the multivariate class (i) where low zHeight and zWeight was observed. Additionally, the “Gradual Increase to High” class shows a strong association with multivariate classes (iii) and (v) which illustrate very different zHeight and zWeight profiles. Multivariate class (iii) describes children with below expected zHeight and expected zWeight scores, while multivariate class (v) illustrates children with expected zHeight and above expected zWeight scores. Thus, while subjects may have a similar relationship between zHeight and zWeight leading to allocation of the same zBMI class, the multivariate zHeight+zWeight approach is able distinguish between these individuals.

Finally, the comparison of agreement between latent trajectories identified when using zWFH and zBMI is presented (Table A10). The strongest associations exist between the zWFH “Sharp Increase to Above Expected” with the zBMI “Gradual Increase to High” classes, the zWFH “Gradual Decrease to Expected” and zBMI “Slight Increase, Slight Decrease to Expected” classes and the zWFH “Gradual Increase to Slightly High” and zBMI “Sharp Increase to High” classes. These associated classes show similar levels at which they settle, however the rate of change up until this point is varied between classes identified using zWFH and zBMI. While the allocation of classes was not entirely in agreement given these two responses, this does suggest that given either zWFH or zBMI as input, the latent trajectories identified across responses may represent similar growth profiles.

### Association with Abnormal Growth

To classify Rapid Weight Gainers (RWG), children were identified who experienced an increase in weight between birth and 9 months that was greater than 0.67 standardized units. The proportion of RWG children within each identified growth trajectory for weight is shown in Table 4.

**Table 4:** Proportion of children that experienced rapid weight gain within the first 9 months of life within the four zWeight growth classes.

|  | **Gradual Decrease to Low** | **Sharp Decrease, Sharp Increase to Expected** | **Gradual Increase; Gradual Decrease to Expected** | **Sharp Increase to High** |
| --- | --- | --- | --- | --- |
| **Normal WG** | 235 (98.7%) | 108 (86.4%) | 206 (43.7%) | 9 (6.8%) |
| **RWG** | 3 (1.3%) | 17 (13.6%) | 265 (56.3%) | 124 (93.2%) |

Most children identified as RWG were allocated to the “Gradual Increase; Gradual Decrease to Expected” and “Sharp Increase to High” zWeight classes, while very few were allocated to the “Gradual Decrease to Low” and “Sharp Decrease, Sharp Increase to Expected” classes (Table 4). 93% of children allocated to the “Sharp Increase to High” class experienced RWG, showing an association with RWG and the high zWeight trajectory. Within the “Gradual Increase; Gradual Decrease to Expected” class, 56% experienced RWG. Thus, this class is almost equally represented by children who experienced RWG, and those who did not, who have settled at a normal weight.

The proportion of those that experienced RWG within other growth trajectories is illustrated within the supplementary materials (Tables A11-A13). Within zBMI, both “Increase to High” trajectories comprised over 50% RWG individuals, this is also seen within the zWFH “Sharp Increase to High” trajectory. Otherwise, the zBMI and zWFH classes do not show a clear association with RWG, which is to be expected as zBMI and zWFH measures consider both zHeight and zWeight profiles. Within the multivariate zHeight+zWeight classes a large percentage of RWG within those allocated to the multivariate classes (iv) and (v) (61% and 81% respectively) occurred, in line with results seen in zWeight.

The number of children that ever experienced some form of abnormal growth, namely stunting (zHeight Score < -2), overweight (zWeight Score > 2) or underweight (zWeight Score < -2), at one or more time points between birth and 5 years were identified. Both “Decrease to Low” (Sharp and Gradual) classes, based on zHeight, show over 50% of individuals classified as stunted at one or more time points (Table 5). Within the “Gradual Increase, Gradual Decrease to Below Expected” class 34% of subjects were classified as stunted at one or more time points, and within the “Sharp Decrease, Sharp Increase to Below Expected” class the largest percentage of 73% of individuals classified as stunted at one or more time points. The largest percentage of children identified as underweight at one or more time points, 50%, were allocated to the “Sharp Decrease, Sharp Increase to Expected” class when considering zWeight (Table 6). Meanwhile, the largest percentage of children identified as overweight at one or more time points, 41%, were allocated to the “Sharp Increase to High” class when considering zWeight (Table 6).

**Table 5:** Proportion of ever stunted children between birth and 5 years as defined by zHeight scores.

|  | **Sharp Decrease to Low** | **Gradual Decrease to Low** | **Sharp Decrease, Sharp Increase to Below Expected** | **Gradual Increase, Gradual Decrease to Below Expected** | **Entire Cohort** |
| --- | --- | --- | --- | --- | --- |
| **Stunted** | 263 (51%) | 61 (55%) | 136 (73%) | 110 (34%) | 570 (50%) |
| **Total** | 516 | 112 | 187 | 321 | 1136 |

**Table 6:** Proportion of children ever underweight or overweight between birth and 5 years as defined by zWeight scores.

|  | **Gradual Decrease to Low** | **Sharp Decrease, Sharp Increase to Expected** | **Gradual Increase; Gradual Decrease to Expected** | **Sharp Increase to High** | **Entire Cohort** |
| --- | --- | --- | --- | --- | --- |
| **Underweight** | 91 (35%) | 75 (50%) | 76 (13%) | 44 (31%) | 286 (25%) |
| **Overweight** | 31 (12%) | 18 (12%) | 77 (13%) | 58 (41%) | 184 (16%) |
| **Total** | 263 | 150 | 584 | 142 | 1139 |

These identified latent profiles thus indicate children with abnormal growth features across the 5-year period rather than at isolated ages.

## Discussion

This paper characterizes the structure and heterogeneity of growth profiles for children from birth to 5 years of age in a LMIC African birth cohort. The use of LCMM allowed identification of distinct classes of growth from birth through 5 years. Three latent classes were identified within zBMI and zWFH; four latent classes were identified within zHeight and zWeight and five latent classes were identified within the multivariate response of zHeight+zWeight. Through these latent classes children were identified who deviated from expected growth patterns and their progression through early childhood was tracked. We identified a clear association between univariate zWeight and zHeight classes and zHeight+zWeight classes and abnormal growth patterns of stunting, underweight, overweight and rapid weight gain.

This is the first study to identify latent growth classes using a joint growth response made up of zHeight and zWeight during childhood. Additionally, the trajectories identified using this joint response were contrasted against those identified using zBMI, zWeight, zHeight and zWFH. The results were internally validated using two specifications of the longitudinal process, through the use of standardised and unstandardised growth responses as well as through repeating the LCMM using internal cross-validation using randomly sampled subsets. Consequently, there is confidence in the latent profiles identified within this report.

Growth responses may be analysed within their observed or standardised formats. The observed, unstandardised approach does not control for prematurity, age or gender. Thus, a key advantage of using standardised growth responses is the generalisability of trajectories, as the impact of gender or gestational age on growth has been removed from the process of latent class identification.

A Cluster Analysis approach could have been used to identify latent groups with respect to growth instead of the LCMM approach. This would have required the definition and analysis of features of a given growth profiles as opposed to the longitudinal growth measurements. Gough et al., used K-means clustering approaches to identify latent trajectories within standardised height (zHeight) scores and identified four classes within this growth response (24). Similarly, Mebrahtu et al., identified three classes within standardised weight (zWeight) scores using the GMM approach (25). Here, within zHeight and zWeight four classes were identified, while zHeight+zWeight lead to the identification of five latent profiles. The classes identified using the multivariate approach were not identical to those found using the univariate approach, and additionally allocation to these classes was not identical. This shows the benefit of considering both the uni- and multivariate approaches when identifying latent growth trajectories. The multivariate approach considers both the change of zHeight and zWeight responses over time that are adjusted for age and sex, with an unrestricted relationship unlike zBMI and zWFH. This also reiterates that the multivariate approach allows the identification of latent trajectories that consider the change of both Height and Weight independently over time. The univariate and multivariate approaches consistently identified a low zHeight class, which may serve as a stunted, or at high risk of stunting class, a low zWeight class (those at greater risk of underweight) and a high zWeight class (those at greater risk of overweight).

Three growth classes were identified within zBMI and zWFH scores between birth and five years. In contrast, Rickman et al. identified four latent classes within zWFH, however they considered zWFH from birth until two years within a Western Kenyan cohort. While they identified an additional class, there is strong agreement between the trajectories identified by Rickman et al. and those within this study; the difference being with an additional low zWFH class identified by Rickman et al. (26). It is possible that cohort differences may explain the difference in profile numbers identified here as Rickman et al., observed an increased risk of suboptimal growth -considering 8 distinct growth measures - within HIV-exposed uninfected children.

Previous studies that make use of latent class analysis to identify groups of children with similar growth trajectories primarily focus on BMI as an indicator of early onset obesity (27,28). Robinson et al., reviewed all publications post 2000 that investigated latent class identification within BMI responses (28). Of the eight studies found, six identified four clear trajectories within BMI during childhood (28). Accordingly, identification of latent classes within BMI or zBMI is seen more frequently in the literature than zWFH. Most commonly, three or four latent classes are identified within zBMI or BMI, however these studies often focus on growth from the age of four onwards (28–31). Wang et al., found dynamic BMI growth patterns (such as BMI catch-up and stable overweight) were more predictive, and thus more informative, than static BMI measurements of cardiovascular structure and function in early childhood (32). Rapid weight gain in early childhood (or catch-up growth) has been associated with an increased risk of overweight, obesity and other chronic diseases during later childhood and adulthood (33–36). Additionally, catch-up growth has been associated with various factors such as altered insulin metabolism (37,38) and hence an increased risk of Type II diabetes (39), increased systolic blood pressure (40), increased risk of coronary heart disease (7) and an increased risk of childhood asthma (41) Various factors may modify the risk of rapid weight gain or obesity, including bottle feeding, shorter gestation age and being firstborn (42,43).

Conventionally Weight-for-Height/Length (zWFH) is used to assess over/under-weight status in children below the age of 2 years, while Body Mass Index (BMI) is used from 2 years onwards (44). Weight-for-Height and standardised BMI (zBMI) scores are calculated using WHO charts; both represent the relationship between Height and Weight. However, zBMI scores are standardised by age while zWFH scores are not (20). Instead, these are created using charts that represent the expected relationship between height and weight between birth and two years or two years and five years of age (45). Aris et al., found that the use of zBMI or zWFH before 2 years did not impact the ability to predict future adiposity or cardiometabolic outcomes in children, suggesting these scores may be equally beneficial.

A clear limitation for the use of zWFH is the lack of age-dependent standardisation, which is resolved when one makes use of zBMI (20,45). However, a shortcoming of the use of BMI to study future risk of obesity, is the fact that BMI does not consistently reflect body composition (46). It has even been suggested that BMI and BMI increase during early childhood is more predictive of lean mass than adiposity during adulthood (47). Body composition, fat mass and fat-free mass may be measured, and subsequent latent classes have been identified within the observations of fat mass and fat-free mass independently (48). However, further research is needed to identify whether this may be a better predictor of obesity in children. Additionally, this requires dedicated machinery that may not be available at many care facilities in Low to Middle Income (LMIC) countries. Here, the trajectories identified within zBMI and zWFH as well as allocation to these classes across these two responses were in some agreement, indicating that there may not be a clear benefit to using zWFH over zBMI. Similarly, Aris et al., considered the association of overweight based on zBMI or zWFH and cardiometabolic risk during the first two years of life and found no clear difference using zBMI or zWFH (45).

Both groups of latent trajectories identified using zBMI and zWFH did not show a strong agreement in allocations when contrasted with multivariate zHeight+zWeight. This illustrates additional information the multivariate zHeight+zWeight approach may add to such analyses. The multivariate zHeight+zWeight approach was able to identify abnormal growth with respect to zHeight, zWeight or both while the zWFH and zBMI approaches focus on zWeight with zHeight as a reference regardless of whether zHeight is classified as normal or abnormal. Thus, the use of multivariate zHeight+zWeight has provided a different perspective to our understanding of growth during childhood.

Remarkably, 31% of overweight children under five live in LMICs, while 72% and 59% of wasted and stunted children under five, respectively, live in LMICs. While the prevalence of stunting and wasting in children is decreasing in Southern Africa, the prevalence of obesity is increasing (1). Fifty percent of subjects within this cohort were identified as stunted at one or more timepoints, in particular the “Sharp Decrease, Sharp Increase to Below Expected” zHeight class. Forty one percent of those allocated to the “Sharp Increase to High” zWeight class were classified as overweight at one or more timepoints. Half of those allocated to the “Sharp Decrease, Sharp Increase to Expected” zWeight class were identified as underweight at one or more timepoints. This illustrates the ability of using identified latent classes as a proxy for those at high risk of stunting, overweight or wasting in a longitudinal setting.

The “Sharp Increase to High” zWeight class identified 93% of RWG children, who are at risk of obesity, and development of non-communicable diseases. Those that experienced RWG make up the majority of children identified within the BMI and WFH “Sharp Increase to High” classes, indicating a higher risk of RWG related adverse effects within these classes. Interestingly the RWG children were allocated to similar classes when considering either zHeight+zWeight or zWeight. Within zHeight+zWeight class (v) 81% of experienced RWG, and hence subjects allocated to this class are at greater risk of RWG associated adverse effects.

Limitations of the analysis described, include the use of a single cohort. However, this cohort comprised of almost 1000 children with numerous repeated growth measurements and high rates of follow-up. Furthermore, this cohort is representative of LMIC child populations with high rates of poverty and infectious diseases, which is a consistently underrepresented population group within latent growth analysis. A potential limitation is the use of WHO and Fenton reference ranges, however these are widely used globally. zHeight and zWeight within this cohort behaved differently from expected, thus, average growth of this population may not be adequately described through the WHO and Fenton growth reference ranges. This would lead to potentially incorrect interpretations of trajectories that may be situated below or above what is expected. The development of a South African specific growth reference would resolve this concern. However, using globally accepted reference ranges allows comparisons with other cohorts and populations globally. As is convention, lying-down length was measured up to two years after which standing height was recorded. It is also well documented that recordings of length often contain more measurement error as it is difficult to keep infants still and equally stretched out during each follow up visit. To minimise this staff were trained repeatedly in growth measurements, a standardised operating procedure was used, and all measurements were done in triplicate. Additionally, due to the study design, observations were acquired more frequently during the first year with observations every six-month interval from one year. However, the first year of life has the most accelerated growth and is the most critical period for development (49), with growth patterns setting a developmental trajectory for life (32,50). This may have impacted the shapes of growth curves identified as there was more information to describe these curves during the first year. The use of a greater degree of smoothing at early ages may have accounted for this impact, this will be considered in future analysis when age ranges are extended.

Longitudinal growth response over time must first be described to ascertain latent growth classes; subsequently latent groupings within such a response may be identified. To describe the longitudinal process a broken-stick model was used. While more flexible growth models such as a cubic-spline, polynomial or more conventional growth models such as the SITAR could have been used none of these approaches would produce as interpretable results as identified here. The broken-stick model allows direct comparison of changes in growth between respective intervals across latent classes. Multiple approaches were considered and compared and did not produce different results. The SITAR approach was unsuccessful given this dataset; this is often the case when using responses with limited visit events.

Future work will focus on comparing latent trajectories with various clustering approaches for multivariate growth measurements. Furthermore, the inclusion of additional growth responses within a multivariate model may allow for the identification of growth profiles that describe the overall growth of individuals throughout childhood. In future we hope to extend this work up until the age of puberty and hope that at this time the increased number of visits will allow the use of the SITAR model for comparison purposes. Additional work will focus on describing determinants of latent growth profiles to identify risk factors for obesity, wasting or stunting. This will also include investigating the association between growth trajectories and childhood or future illness, hence identifying possible areas of intervention to promote optimal growth in children. Here the understanding of how Weight and Height may change throughout childhood has been broadened. With the identification of these distinct growth trajectories, one may be able to explore whether these specific profiles may serve as an indicator of early onset illness.

### Ethics Approval

The study was approved by the faculty of Health Sciences, Human Research Ethics Committee, University of Cape Town (401/2009), Stellenbosch University (N12/02/0002) and the Western Cape Provincial Health Research committee (2011RP45).

### Data Availability

Researchers who are interested in datasets or collaborations can contact the PI, Heather Zar with a concept note outlining the request. More information can be found on our website (http://www.paediatrics.uct.ac.za/scah/dclhs).

### Supplementary Data

Supplementary data are available.

### Author contributions

HZ designed and oversaw the Drakenstein study and obtained funding. FL designed and supervised the analytical strategy and helped to interpret the findings. ML prepared the data for analysis and assisted with analytic aspects. NVB primarily drafted the manuscript and performed the statistical analysis. LG oversaw nutritional aspects of the study and provided clinical expertise. MB assisted with data aspects and operational oversight. All authors contributed to editing of the final manuscript.

## Funding

This study was funded by Bill and Melinda Gates Foundation (OPP1017641; OPP1017579), the NIH, USA (U54HG009824, U01AI110466], South African Medical Research Council, and the University of Cape Town (UCT). NVB was supported through a fellowship to UCT from the Developing Emerging African Leaders (DEAL) programme by The Carnegie Corporation of New York and a Departmental Fellowship from the Department of Statistical Sciences, UCT.

## Acknowledgments

The authors thank the study and clinical staff at Paarl Hospital, Mbekweni and TC Newman clinics, as well as the CEO of Paarl Hospital, Dr. Kruger, and the Western Cape Health Department for their support of the study. The authors thank the families and children who participated in this study.

## Conflict of Interests

None declared.

## Notes

### Competing Interest Statement

The authors have declared no competing interest.

