## Supplementary Materials for "Latent Classes of Anthropometric Growth in Early Childhood Using Uni- and Multivariate approaches in a South African Birth Cohort"

Part 1. Study Design

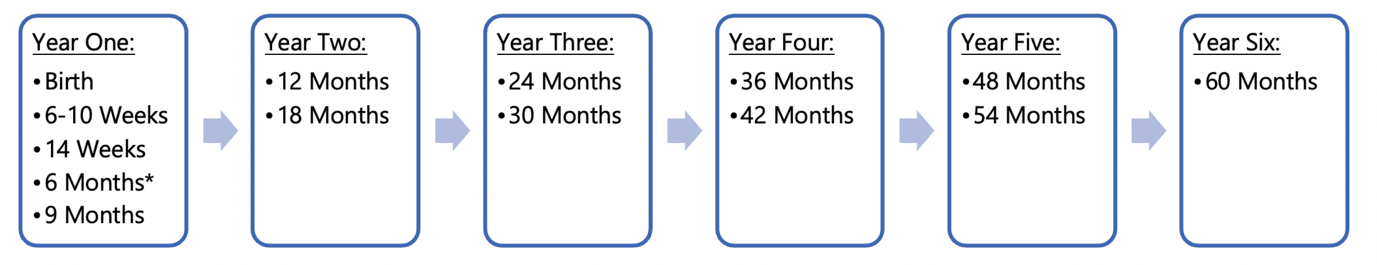
Figure A1: Time-line of follow-up visits for subjects within the DCHS.

Height, Weight, Head Circumference, BMI and WFH recorded at all visits.

* the age from which TRI and MUAC observations have been recorded.

Table A1: The mean growth response (with standard deviations) and number of observations at each follow up visit from Birth until 60 months of age.

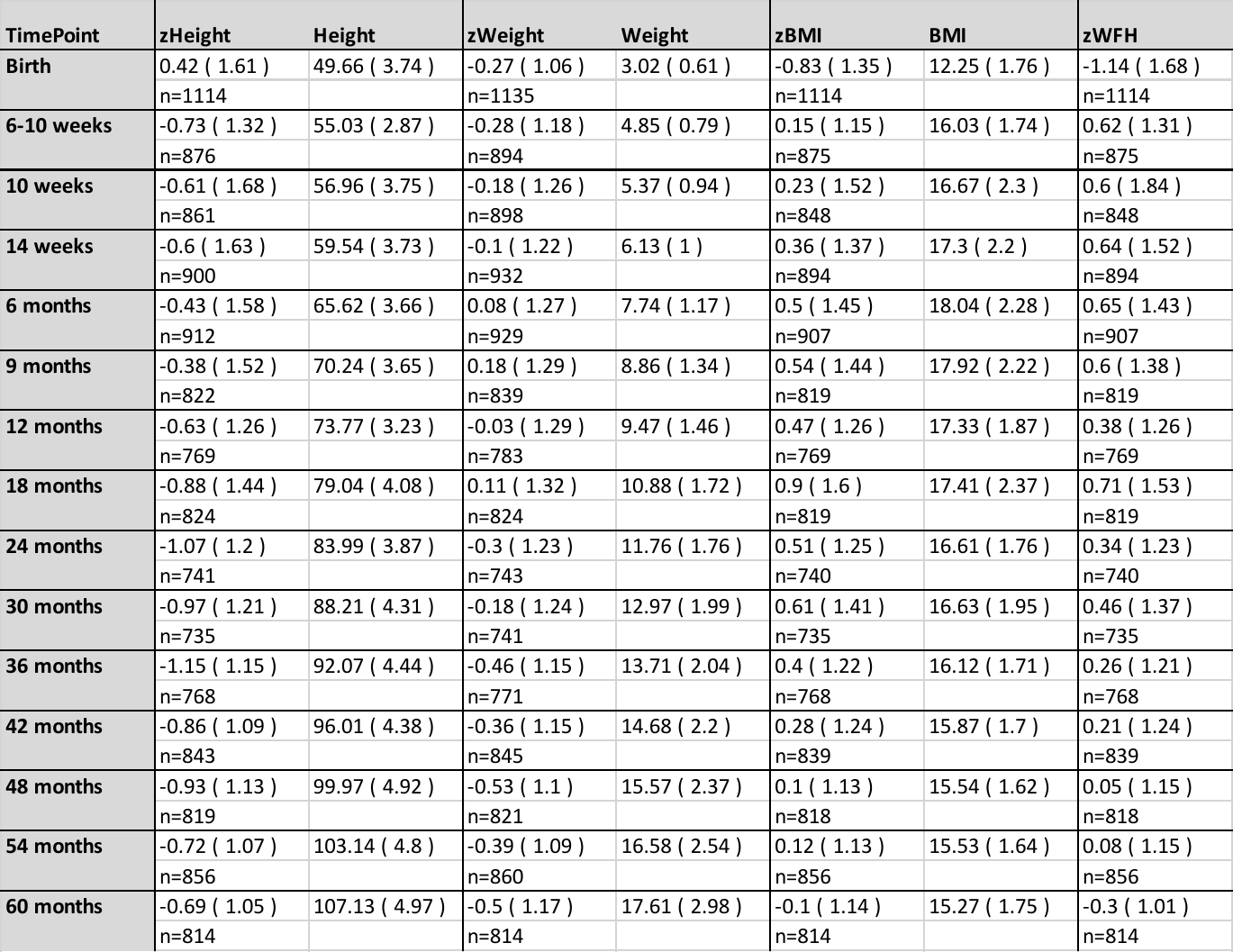

Table A2: Summary of Maternal, Child and Socio-Economic Characteristics

| Maternal Characteristics at Birth (n=1137) | | Total Cohort | Completed (n=981) | Excluded due to LTFU or Death (n=162) |
| --- | --- | --- | --- | --- |
| Mean Maternal Age at Birth (years) |  | 26.9 (sd= 5.72) | 27.1 (sd= 5.75) | 26.1 (sd=5.44) |
| HIV Infected |  | 248 (21.7%) | 216 (22.0%) | 31 (19.8%) |
| Primigravida (n=1135) |  | 392 (34.5%) | 328 (33.5%) | 64 (39.5%) |
| Highest Education Level Attained | Primary | 86 (7.6%) | 72 (7.4%) | 12 (7.5%) |
| (n=1135) | Some Secondary | 609 (53.7%) | 542 (55.2%) | 67 (41.9%) |
|  | Completed Secondary | 375 (33.0%) | 312 (31.8%) | 63 (39.4%) |
|  | Any Tertiary | 73 (6.4%) | 55 (5.6%) | 18 (11.3%) |
| Self-reported Antenatal Smoking (n=1130) |  | 262 (23.2%) | 237 (24.2%) | 25 (15.6%) |
| Prenatal Alcohol Exposure (n=1061) |  | 137 (12.9%) | 121 (12.3%) | 16 (12.5%) |
| Socio-Economic Characteristics at Birth (n=1143) |  |  |  |  |
| Average household income per month | <1000ZAR | 431 (37.7%) | 344 (35.1%) | 42 (26.1%) |
| (n=1142) | 1000-5000ZAR | 557 (48.8%) | 515 (52.5%) | 81 (50.3%) |
|  | >5000ZAR | 155 (13.6%) | 122 (12.4%) | 38 (23.6%) |
| Food Insecure (antenatally; n=1004) |  | 279 (27.8%) | 251 (25.6%) | 28 (23.3%) |
| Child Demographics (n=1143) | |  |  |  |
| Sex | Male | 586 (51.3%) | 501 (51.1%) | 85 (52.5%) |
| Birth Weight (kg) |  | 3.02 (sd=0.61) | 3.30 (sd=0.58) | 2.95 (sd=0.73) |
| Birth Height (cm) |  | 49.66 (sd=3.74) | 49.70 (sd=3.62) | 49.41 (sd=4.40) |
| Gestation | Preterm (< 37 weeks) | 191 (16.7%) | 155 (15.8%) | 37 (22.8%) |
|  | Late Preterm (34-37 weeks) | 128 (11.2%) | 109 (11.1%) | 20 (12.3%) |
|  | Median Gestational Age (weeks) | 39 (sd=2.66) | 39 (sd=2.46) | 39 (sd=3.63) |
| Breast Feeding | Initiated (n=1066) | 868 (81.4%) | 802 (81.8%) | 66 (73.3%) |
|  | Exclusive Up To 6 Months (n=1030) | 72 (7.0%) | 70 (7.1%) | 2 (2.2%) |
| HIV | Unexposed | 894 (78.2%) | 765 (78.0%) | 130 (80.2%) |
|  | Exposed-Uninfected | 247 (21.6%) | 216 (22.0%) | 32 (19.8%) |
|  | Infected | 2 (0.002%) | 2 (0.002%) | 0 (0%) |

Part 2: Extended Statistical Methods

The final form of the mixed effect model for each of the responses takes the following structure:

$$y_{ij}=\beta_{0}+\beta_{1}x_{1ij}+b_{1}\left( x_{1ij}-l_{1} \right)_{+}+b_{2}\left( x_{1ij}-l_{2} \right)_{+}+\ldots+b_{n}\left( x_{1ij}-l_{n} \right)_{+}+\mu_{i}+\epsilon_{ij}$$

where,

$y_{ij}$ refers to the growth response measured for individual i at time point j,

$x_{1ij}$ refers to the age of individual i at time point j,

$\mu_{i}$ captures the *subject specific random effect,*

$\epsilon_{ij}$ captures the *within-subject error,*

$l_{1 to n}$ refer to knot locations with n included knots.

The linear mixed model assumes all subjects were sampled from the same population and thus can be represented using the same population-level relationship between the response and covariates. The latent class mixed model (LCMM) however assumes that the subjects have been sampled from K groups (or populations) and follow different latent profiles (Proust-Lima et al., 2015).

The LCMM method makes use of three models: a linear mixed effect model – the structural model (1), a measurement model – which relates the latent process to the observations (2) and a multinomial logistic model - which describes class membership (3).

(1) the structural model,

$\Lambda_{i}\left( t \right)|_{c_{i}=k}=X_{L_{1}i}\left( t \right)^{T}\beta+X_{L_{2}i}\left( t \right)^{T}\nu_{k}+Z_{i}\left( t \right)^{T}u_{ik}$;	where

$\Lambda_{i}\left( t \right)$ refers to the latent process for subject i at timepoint t,

$c_{i}=k$ if subject belongs to latent class $k$,

and assume population composed of K underlying latent classes characterised by K, mean profiles of trajectories with latent class variable,

$\beta$ describes the effects of the population level covariates, $X_{L_{1}i}$,

within this study this refers to the time component and hence $X_{L_{1}i}\left( t \right)^{T}\beta$ refers to the piecewise-linear spline specification previously described,

$\nu_{k}$ describes the effects of group level covariates, $X_{L_{2}i}$,

$u_{ik}$ describes the subject specific profiles within this model,

thus, capturing the repeated measures nature of this data. which describes the growth responses over time and allows for repeated measures data.

(2) the measurement model,

$Y_{ij}=H\left( \Lambda_{i}\left( t_{ij} \right)+\epsilon_{ij};\eta\right)$	where

$Y_{ij}$denotes the response for subject $i$ at occasion $j$,

$H$ is a parameterized (with $\eta$) link function,

$\epsilon_{ij}$ captures within-subject error,

allowing for different types of response variables and a nonlinear relationship, which relates the latent process to the observed responses.

(3) the multinomial logistic model,

$\pi_{ik}=P\left( c_{i}=k|X_{ci} \right)=\frac{e^{\xi_{0k}+X_{ci}^{T}\xi_{1k}}}{\sum_{l=1}^{K} e^{\xi_{0l}+X_{ci}^{T}\xi_{1l}}}$

where:

$X_{ci}$ refers to covariates that are included as control variables within the class allocation process,

$\xi_{0k}$ is the intercept for class k,

$\xi_{1k}$is the vector of class-specific parameters associated with the time-independent covariates $X_{ci}$,

describing group membership, based on a multinomial logistic model from which subjects are allocated to the class with the greatest resulting probability (Proust-Lima et al., 2015)*.*

Within this study we did not consider the effect of additional covariates and hence this is simplified to the following:

(1) the structural model,

$\Lambda_{i}\left( t \right)|_{c_{i}=k}=X_{L_{1}i}\left( t \right)^{T}\beta+Z_{i}\left( t \right)^{T}u_{ik}$;

(2) the measurement model,

$Y_{ij}=H\left( \Lambda_{i}\left( t_{ij} \right)+\epsilon_{ij};\eta\right)$

(3) the multinomial logistic model,

$\pi_{ik}=P\left( c_{i}=k|X_{ci} \right)=\frac{e^{\xi_{0k}}}{\sum_{l=1}^{K} e^{\xi_{0l}}}$

The LCMM model can be extended to model multiple responses simultaneously by fitting multivariate mixed models and multivariate latent class mixed models for multivariate longitudinal response variables. In this instance, the models previously described are extended to allow observations from *m* responses, from i individuals at j timepoints. This is described in greater detail within Figure 2.

Part 3: Latent Growth Response Analysis Results

*3.1: Longitudinal Growth Responses*

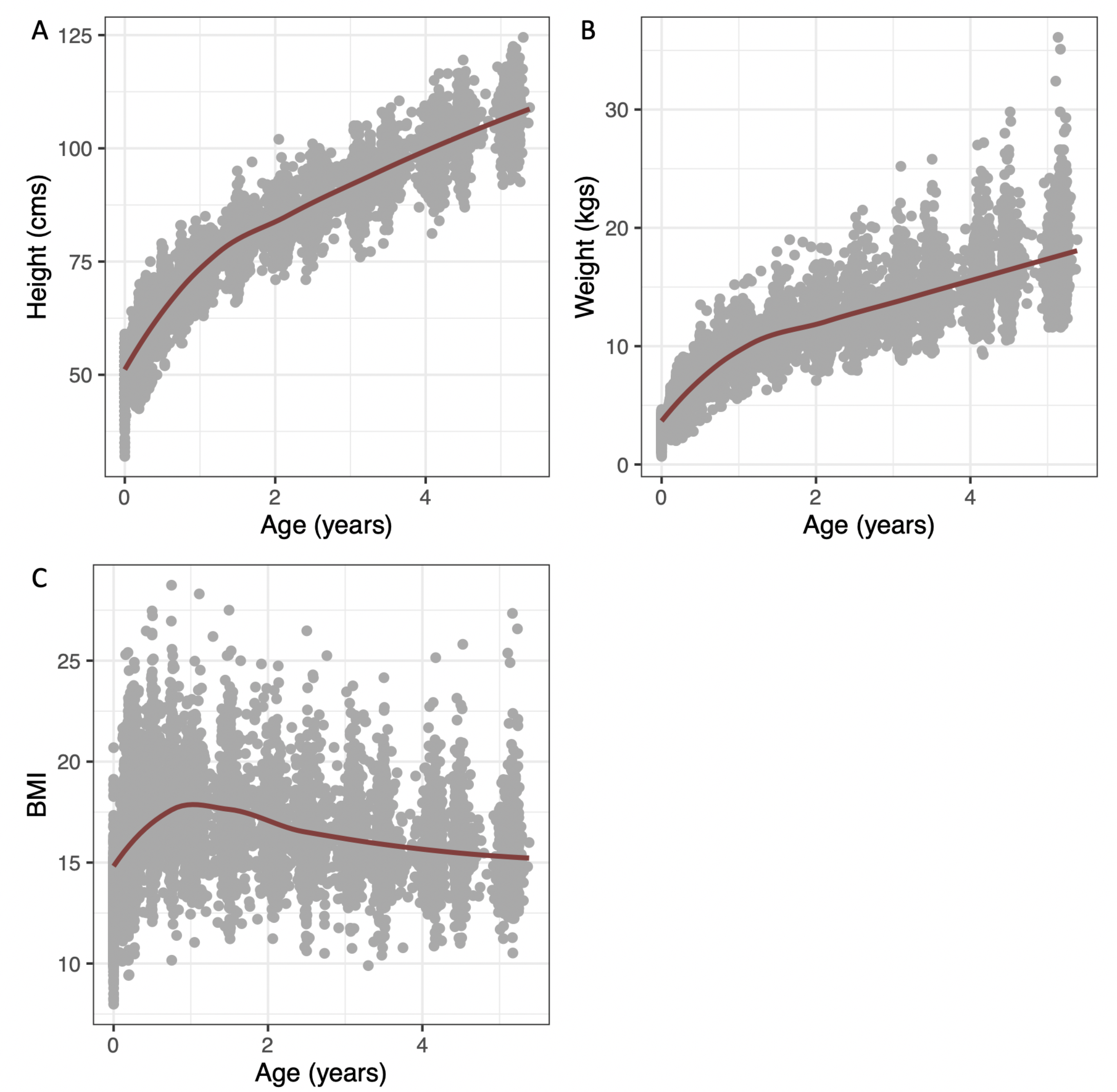

Figure A2: Observed growth measurements over time with a smoothed average indicated with a burgundy line for A) Height, B) Weight and C) BMI.

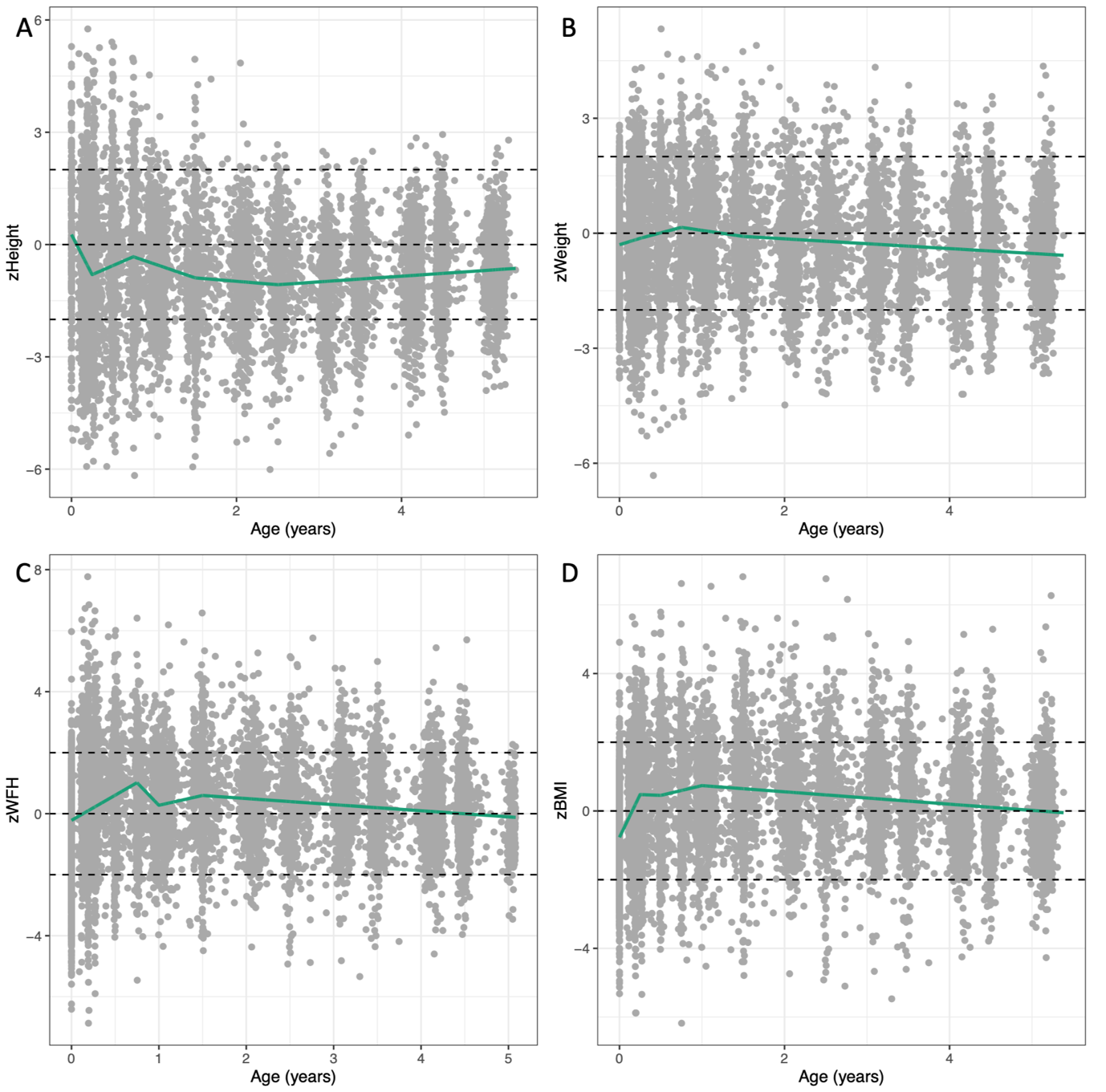

Figure A3: Standardised growth measurements over time with the average trajectory described through a piecewise-linear spline indicated with a green line for A) zHeight, B) zWeight, C) zWFH and D) zBMI.

*3.2: Average Trajectory given Increased Knot Locations*

Table A3: Knot locations for the linear mixed effect models fit using a piecewise linear spline to describe the association between the respective growth measures and age.

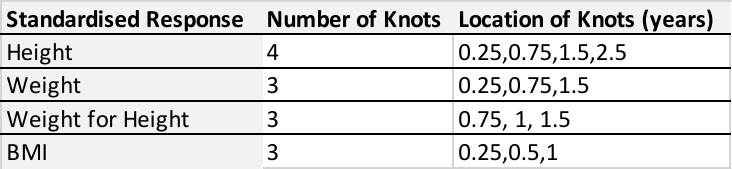

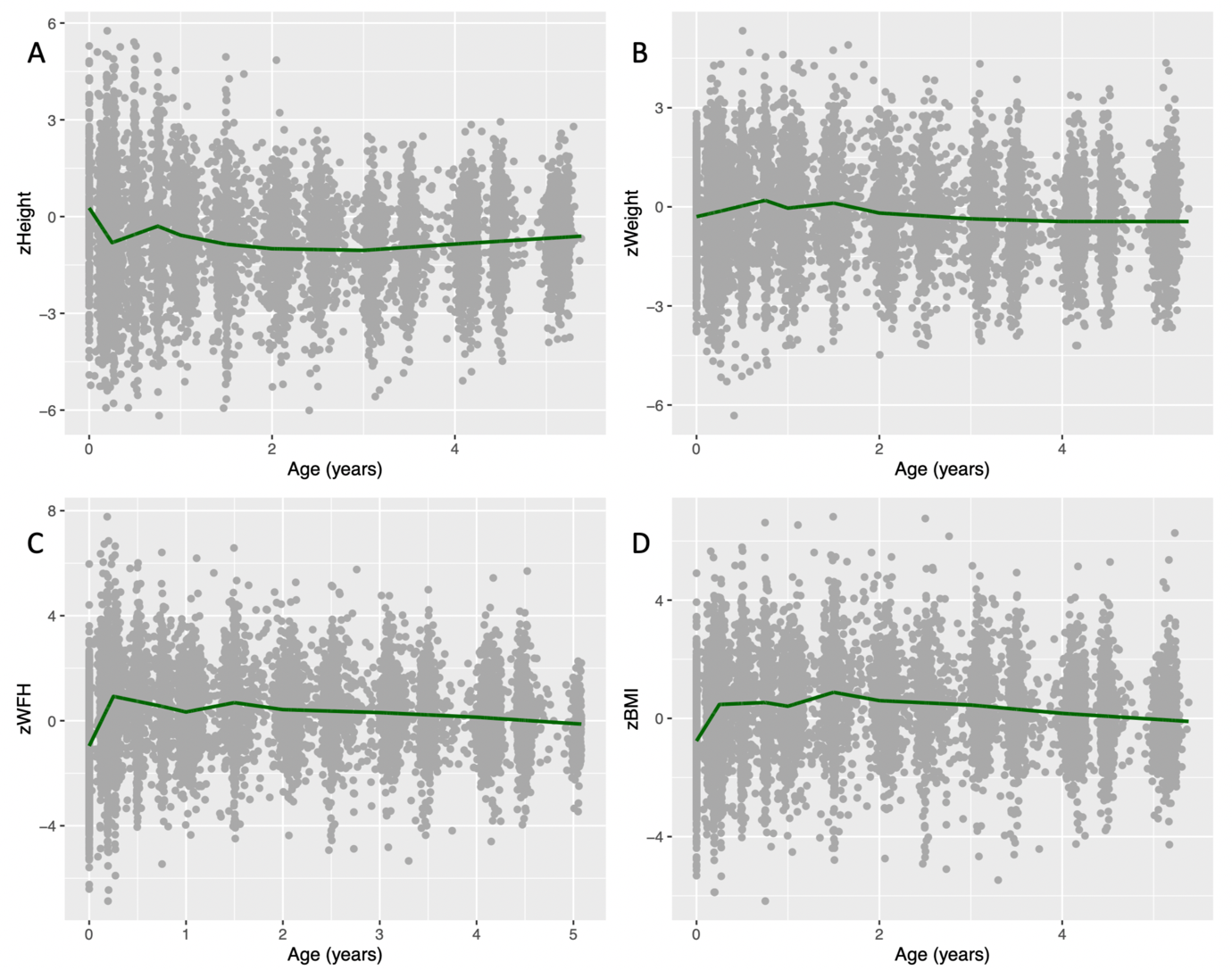

Figure A4: Standardised growth measurements over time with the average trajectory described through a piecewise-linear spline indicated with a green line for A) zHeight, B) zWeight, C) zWFH and D) zBMI given additional knots placed at timepoints (0.25,0.75,1,1.5,2,3,4).

3.3: Stability Testing
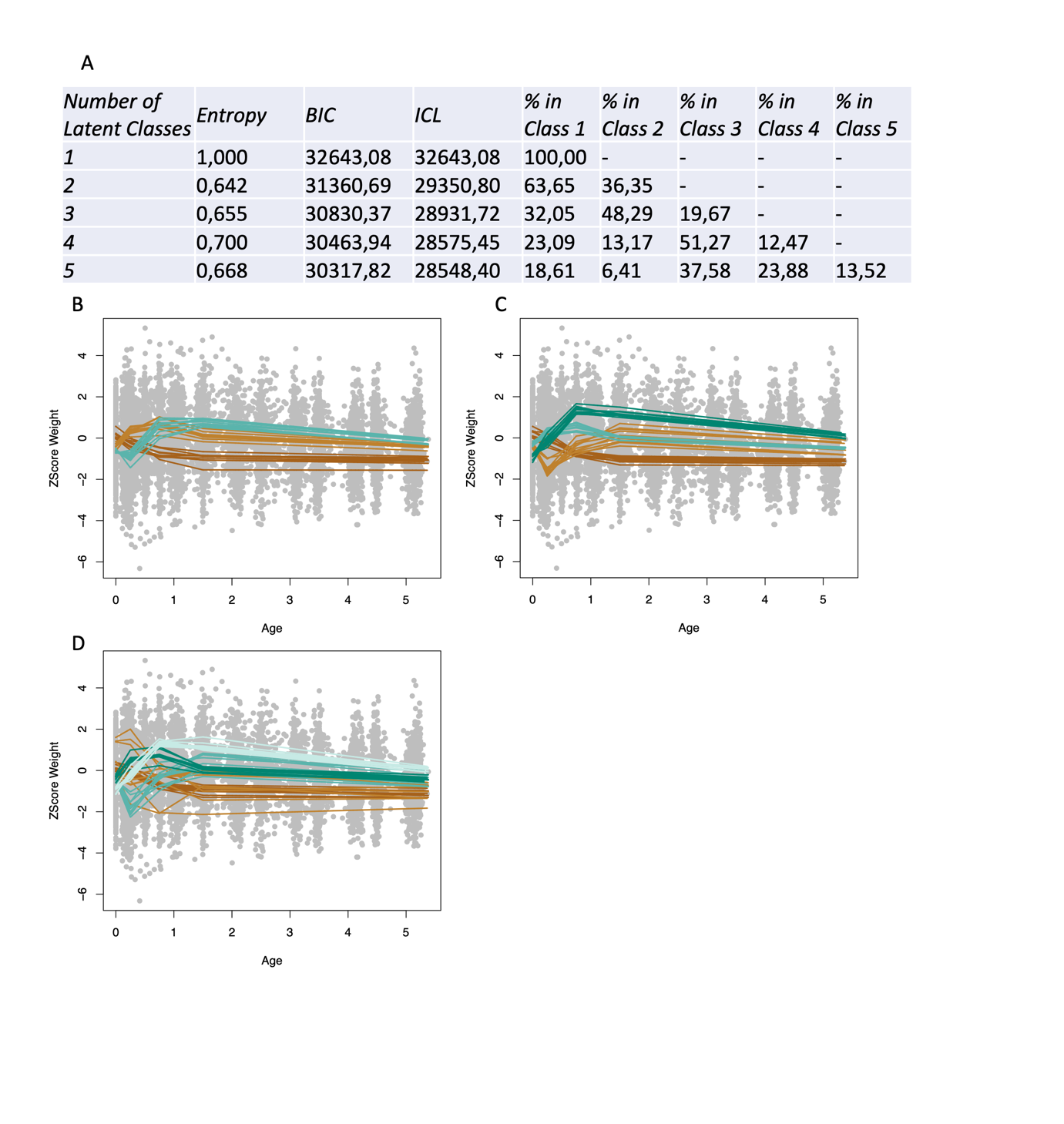
 Figure A5: Information used to choose the appropriate value for k, the number of latent classes within standardised Weight. A) Fit statistics for k=(1:5). Profiles of LCMM Classes identified within standardised Weight using a randomly selected 50% of subjects, repeated 10 times for B) k=3, C) k=4 and D) k=5.

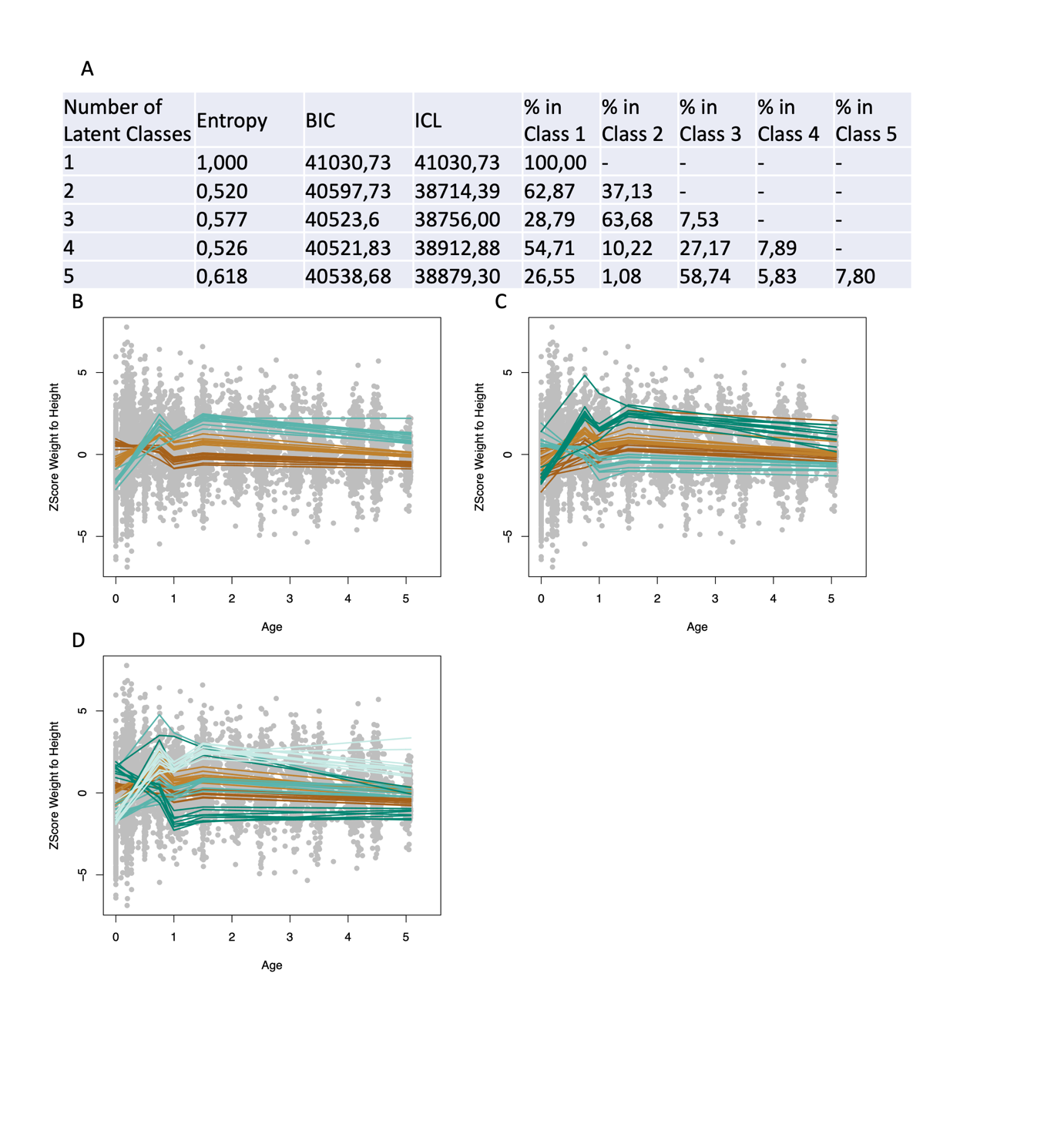
Figure A6: Information used to choose the appropriate value for k, the number of latent classes within standardised WFH A) Fit statistics for k=(1:5). Profiles of LCMM Classes identified within standardised WFH using a randomly selected 50% of subjects, repeated 10 times for B) k=3, C) k=4 and D) k=5.

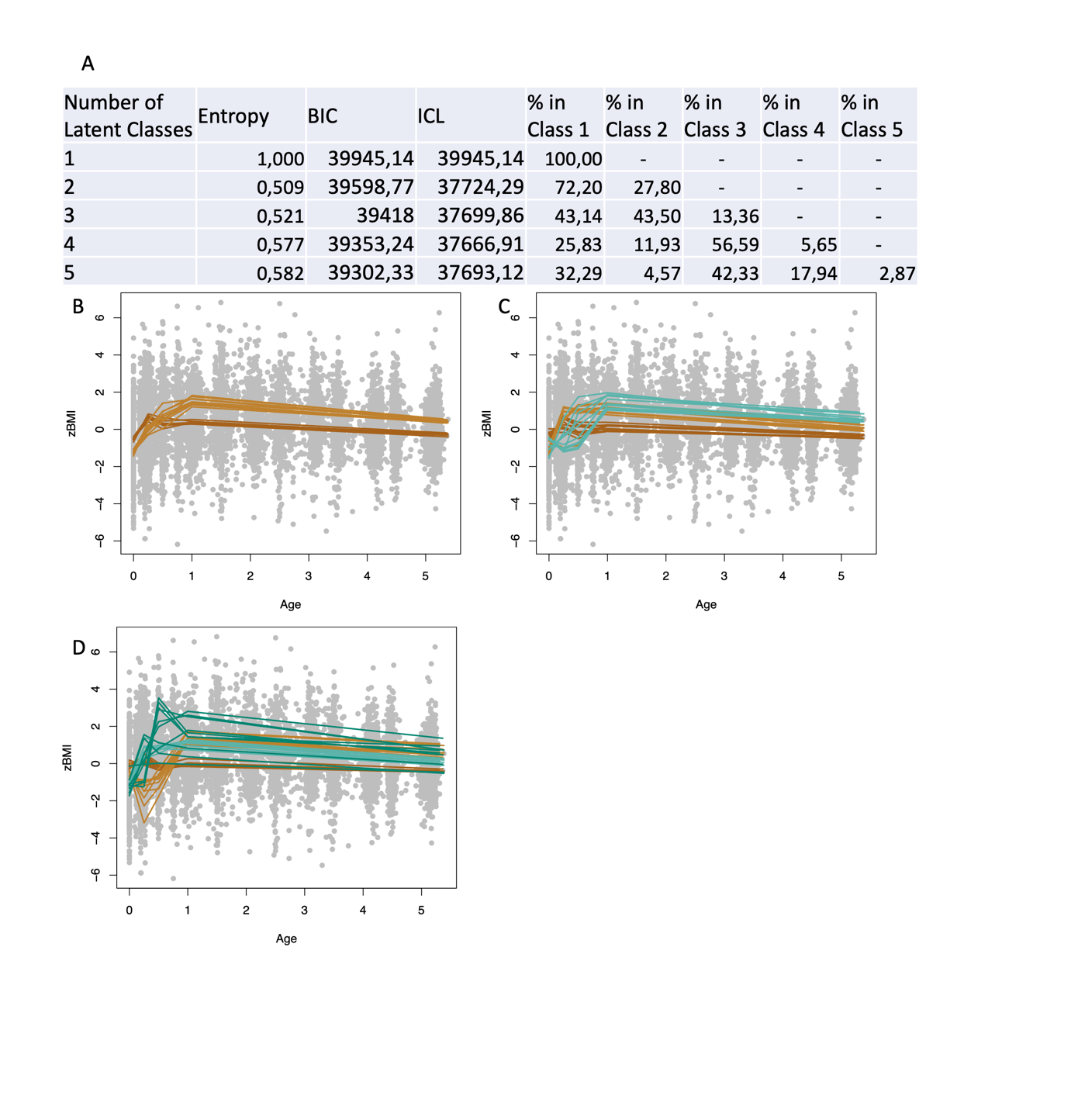
Figure A7: Information used to choose the appropriate value for k, the number of latent classes within standardised BMI A) Fit statistics for k=(1:5). Profiles of LCMM Classes identified within standardised BMI using a randomly selected 50% of subjects, repeated 10 times for B) k=3, C) k=4 and D) k=5.

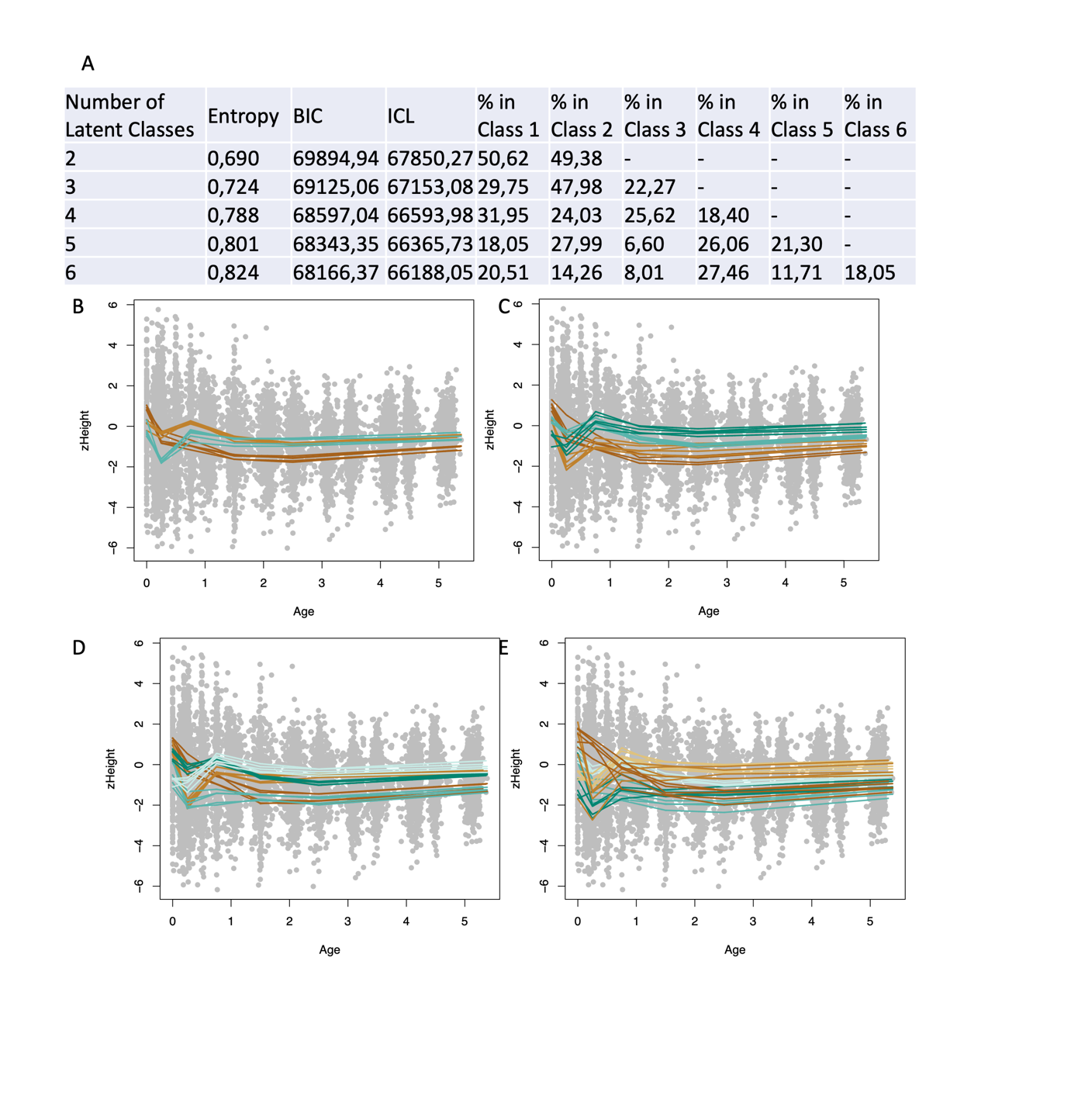
Figure A8: Information used to choose the appropriate value for k, the number of latent classes within zHeight + zWeight. Profiles of LCMM Classes identified within zHeight + zWeight using a randomly selected 50% of subjects, repeated 10 times for A) k=3, B) k=4, C) k=5 and D) k=6 illustrated using zHeight.

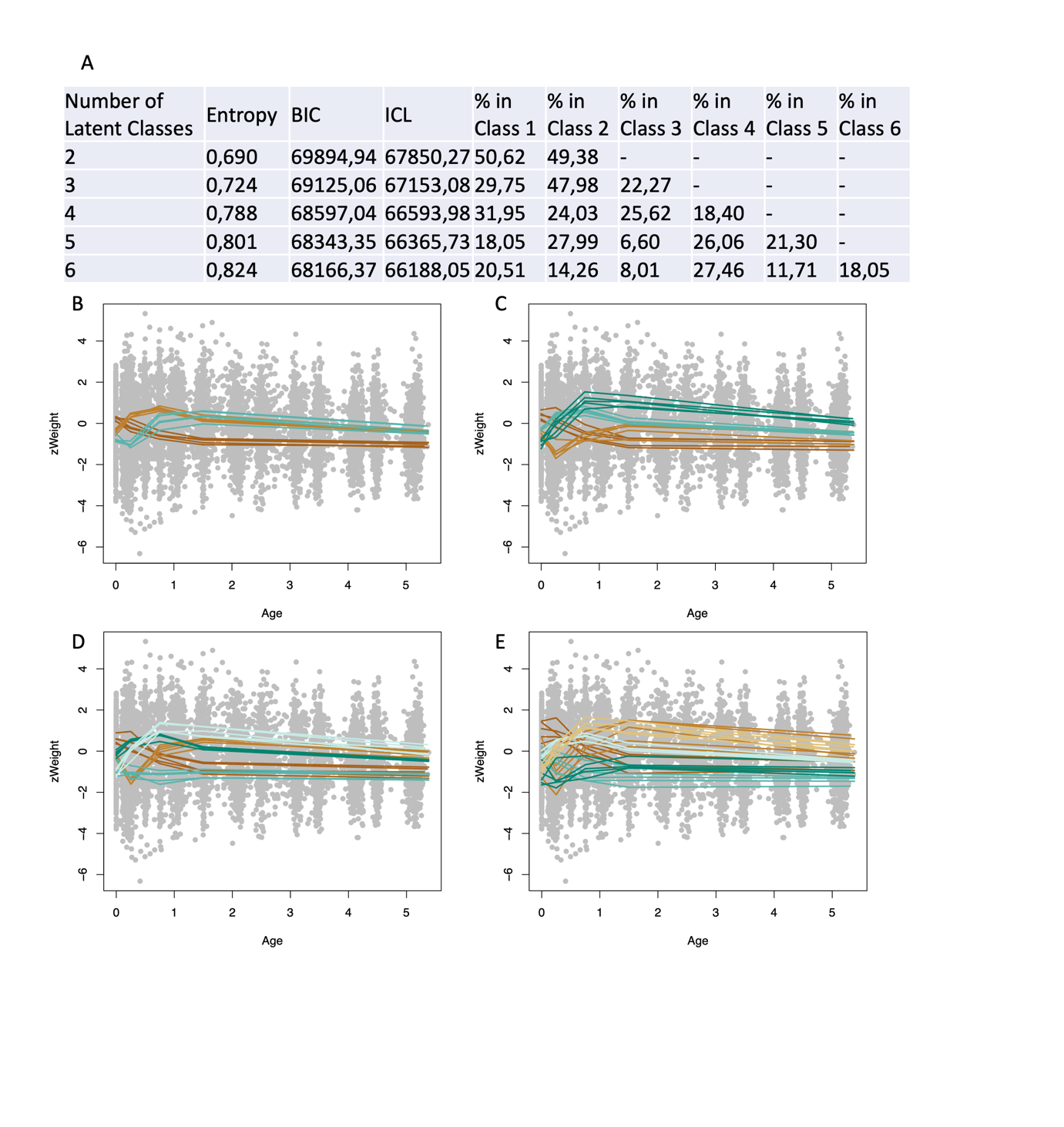
Figure A9: Information used to choose the appropriate value for k, the number of latent classes within zHeight + zWeight A) Fit statistics for k=(1:6). Profiles of LCMM Classes identified within zHeight + zWeight using a randomly selected 50% of subjects, repeated 10 times for B) k=3, C) k=4, D) k=5 and E) k=6, illustrated using zWeight.

*3.4: Stability Testing given Increased Knot Locations*
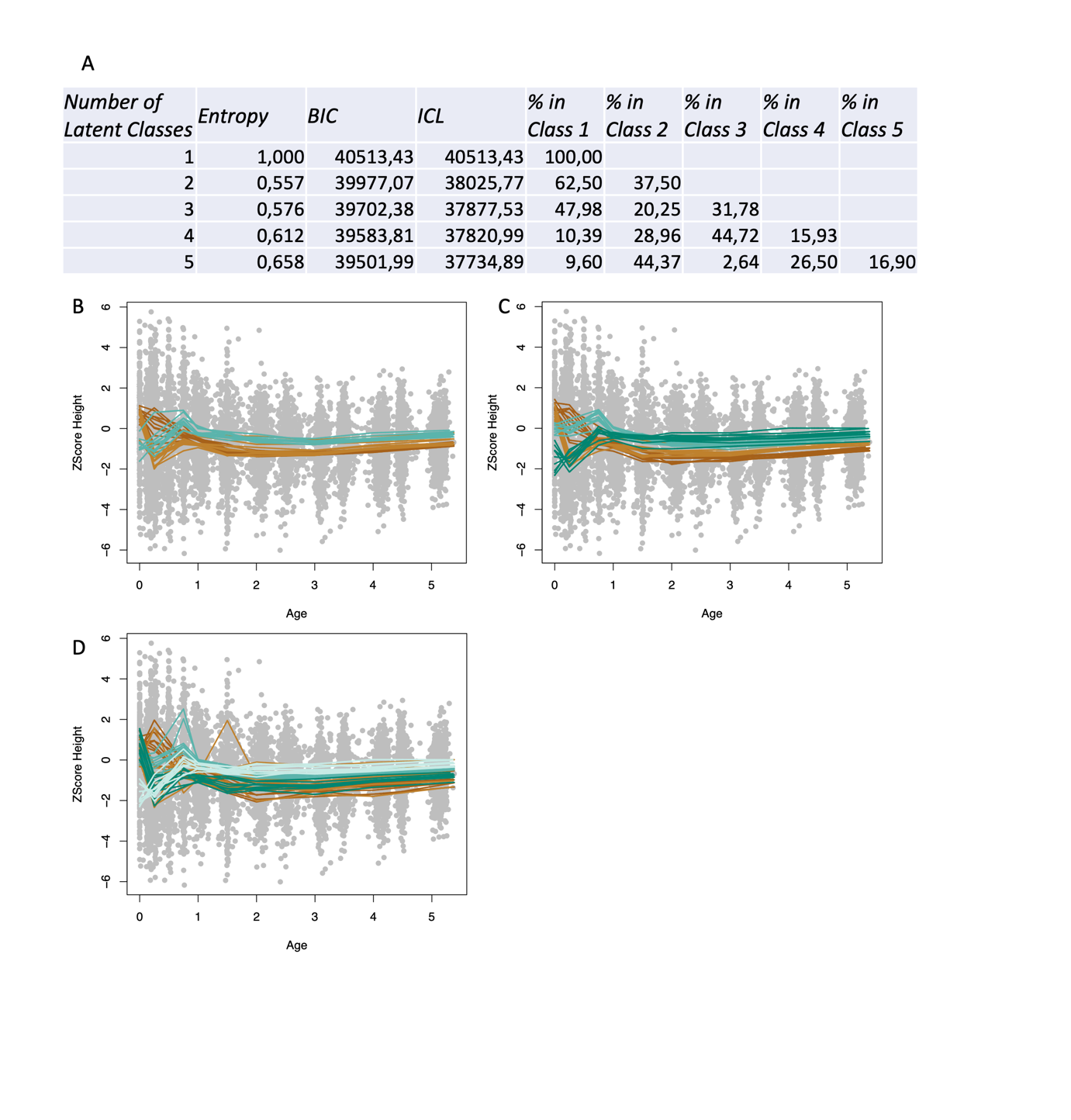
Figure A10: Information used to choose the appropriate value for k, the number of latent classes within standardised Height, given a piecewise linear spline model specification with knots places at (0.25,0.75,1,1.5,2,3,4). A) Fit statistics for k=(1:5). Profiles of LCMM Classes identified within standardised Height using a randomly selected 50% of subjects, repeated 10 times for B) k=3, C) k=4 and D) k=5.

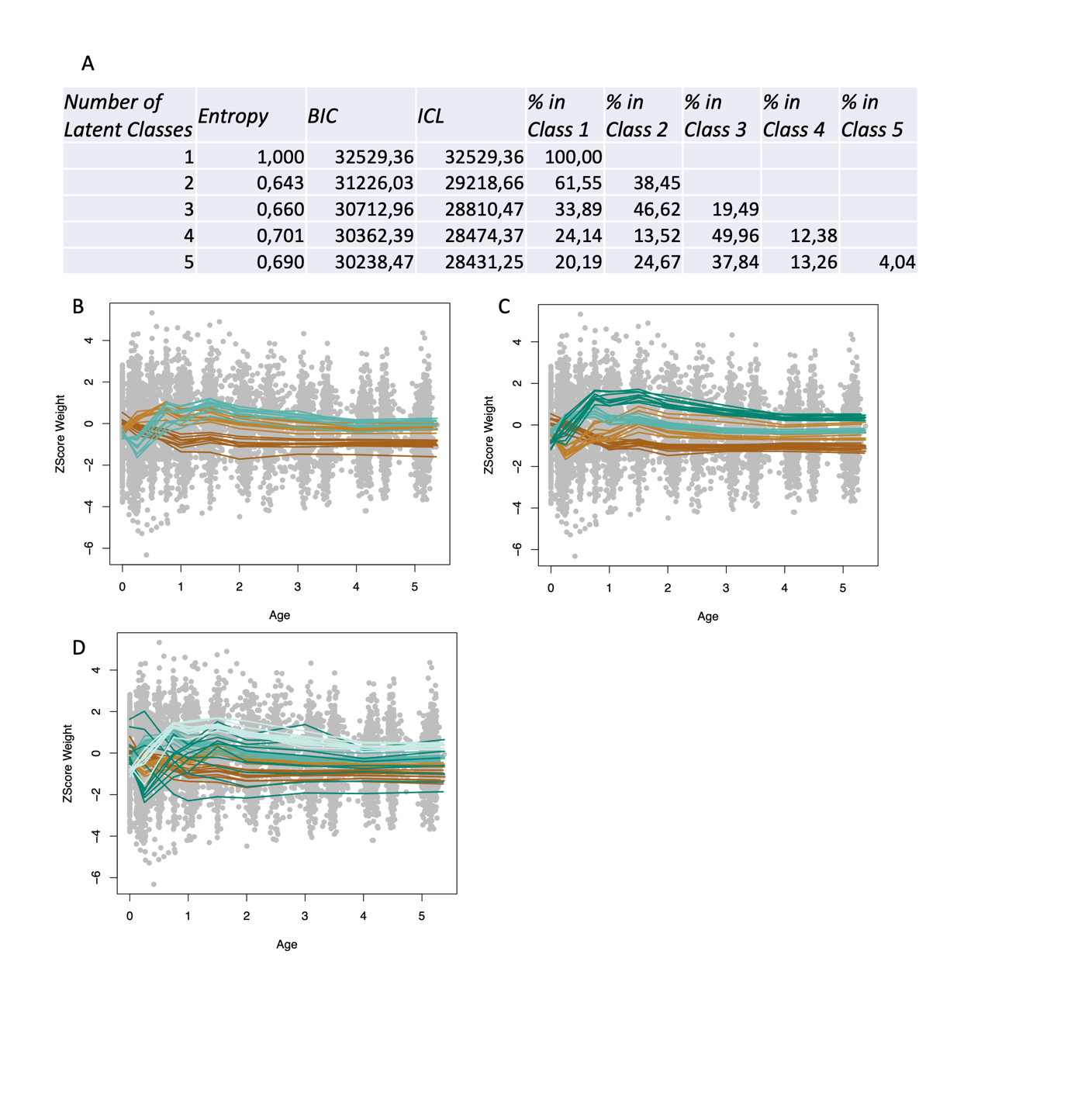
Figure A11: Information used to choose the appropriate value for k, the number of latent classes within standardised Weight, given a piecewise linear spline model specification with knots places at (0.25,0.75,1,1.5,2,3,4). A) Fit statistics for k=(1:5). Profiles of LCMM Classes identified within standardised Weight using a randomly selected 50% of subjects, repeated 10 times for B) k=3, C) k=4 and D) k=5.

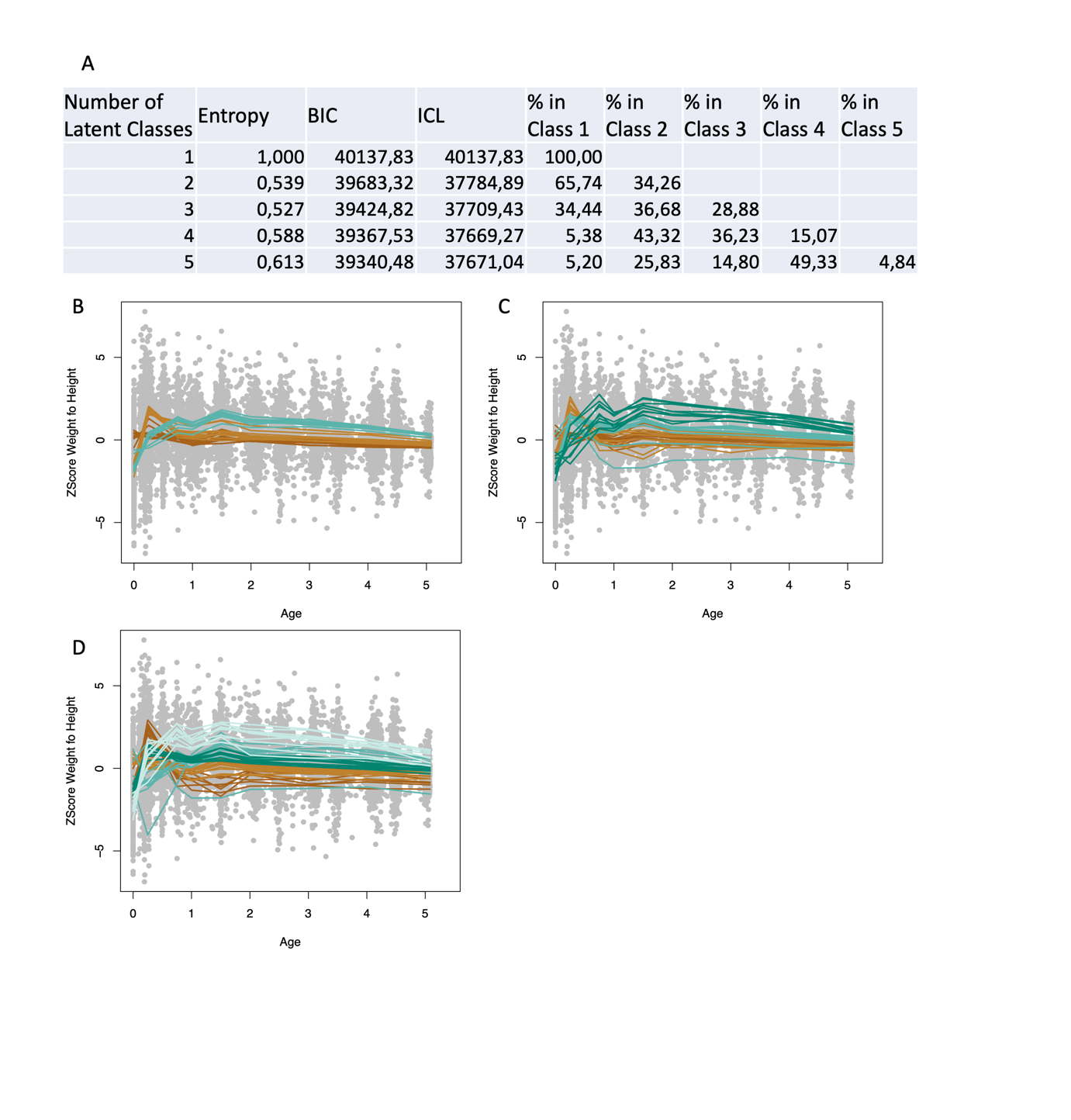
Figure A12: Information used to choose the appropriate value for k, the number of latent classes within standardised WFH, given a piecewise linear spline model specification with knots places at (0.25,0.75,1,1.5,2,3,4). A) Fit statistics for k=(1:5). Profiles of LCMM Classes identified within standardised WFH using a randomly selected 50% of subjects, repeated 10 times for B) k=3, C) k=4 and D) k=5.

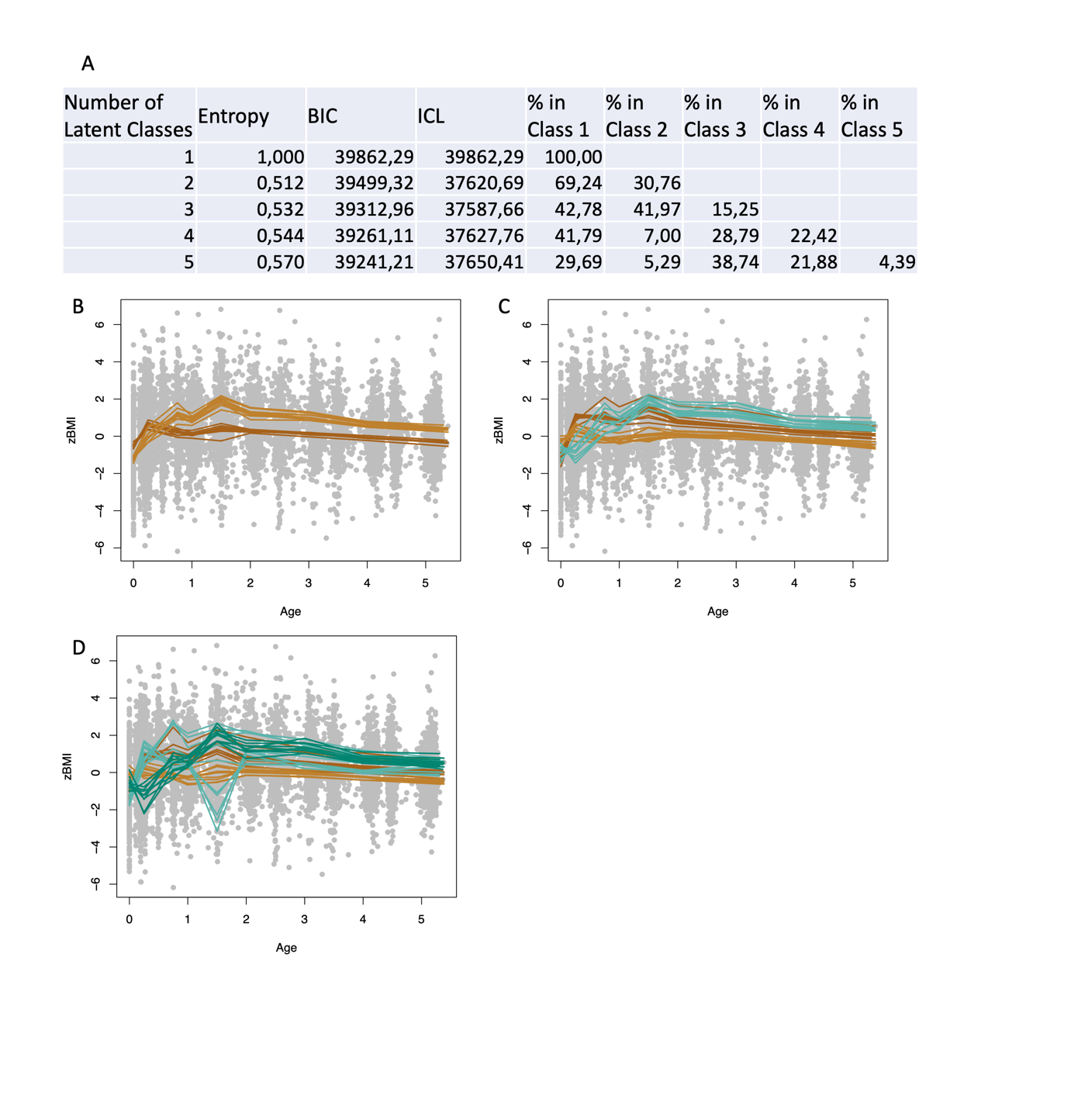
Figure A13: Information used to choose the appropriate value for k, the number of latent classes within standardised BMI, given a piecewise linear spline model specification with knots places at (0.25,0.75,1,1.5,2,3,4). A) Fit statistics for k=(1:5). Profiles of LCMM Classes identified within standardised BMI using a randomly selected 50% of subjects, repeated 10 times for B) k=3, C) k=4 and D) k=5.

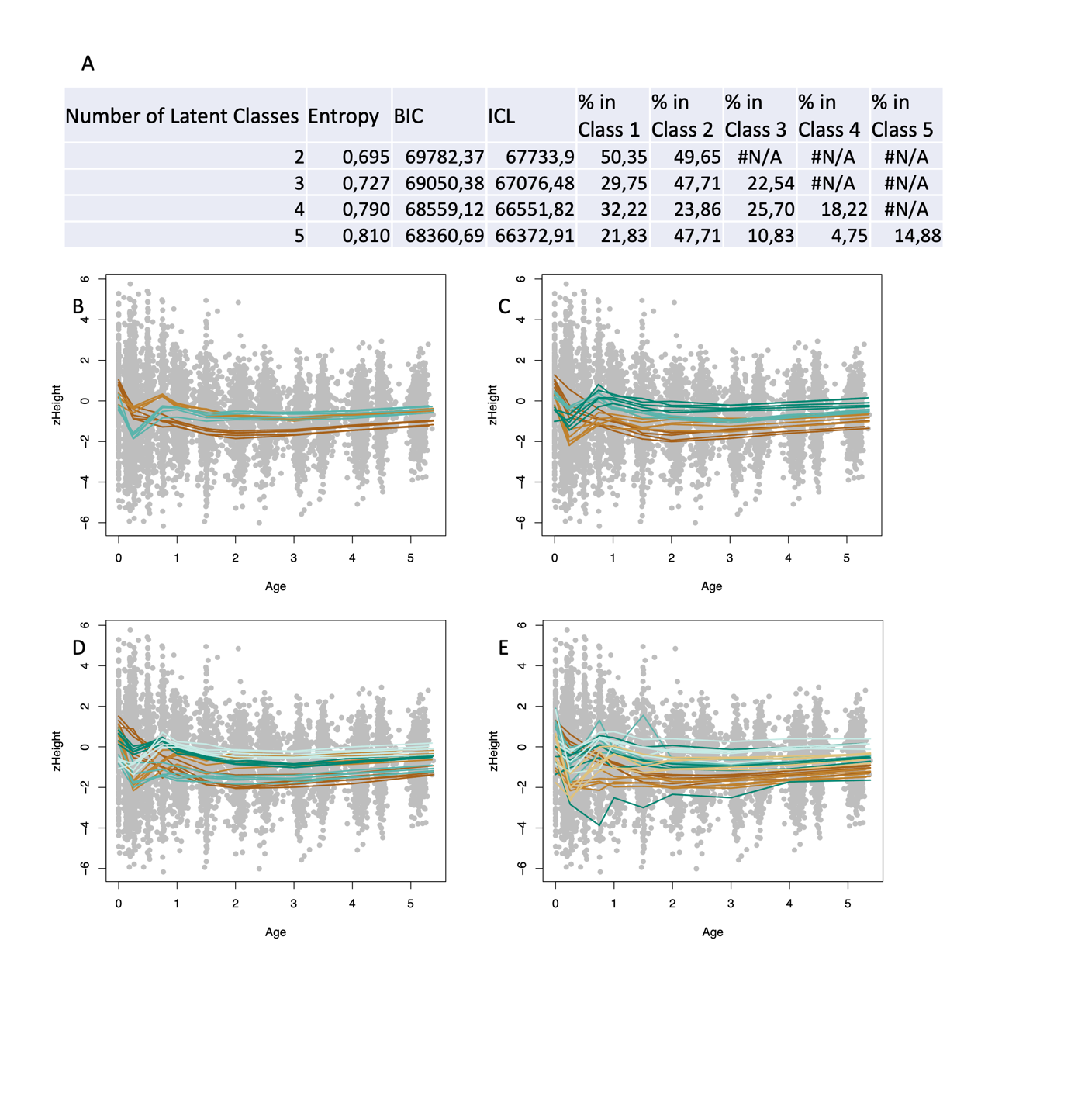
Figure A14: Information used to choose the appropriate value for k, the number of latent classes within zHeight + zWeight, given a piecewise linear spline model specification with knots places at (0.25,0.75,1,1.5,2,3,4). Profiles of LCMM Classes identified within zHeight + zWeight using a randomly selected 50% of subjects, repeated 10 times for A) k=3, B) k=4, C) k=5 and D) k=6 illustrated using zHeight.

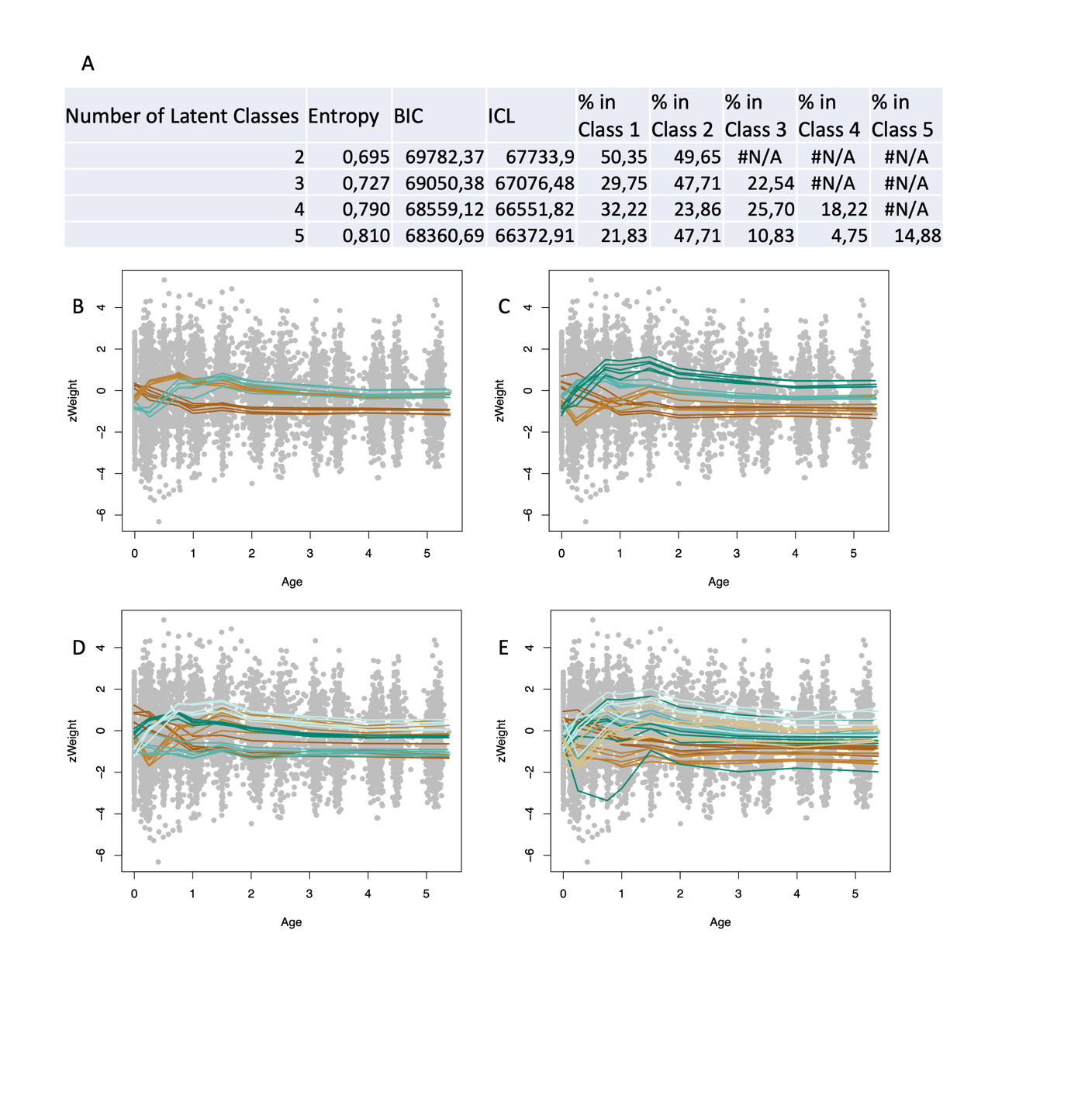
Figure A15: Information used to choose the appropriate value for k, the number of latent classes within zHeight + zWeight, given a piecewise linear spline model specification with knots places at (0.25,0.75,1,1.5,2,3,4). A) Fit statistics for k=(1:6). Profiles of LCMM Classes identified within zHeight + zWeight using a randomly selected 50% of subjects, repeated 10 times for B) k=3, C) k=4, D) k=5 and E) k=6, illustrated using zWeight.

*3.5: Final Model Estimates*

Table A4: Fixed Effect Estimates from the Measurement model within the LCMM framework given the zHeight response.

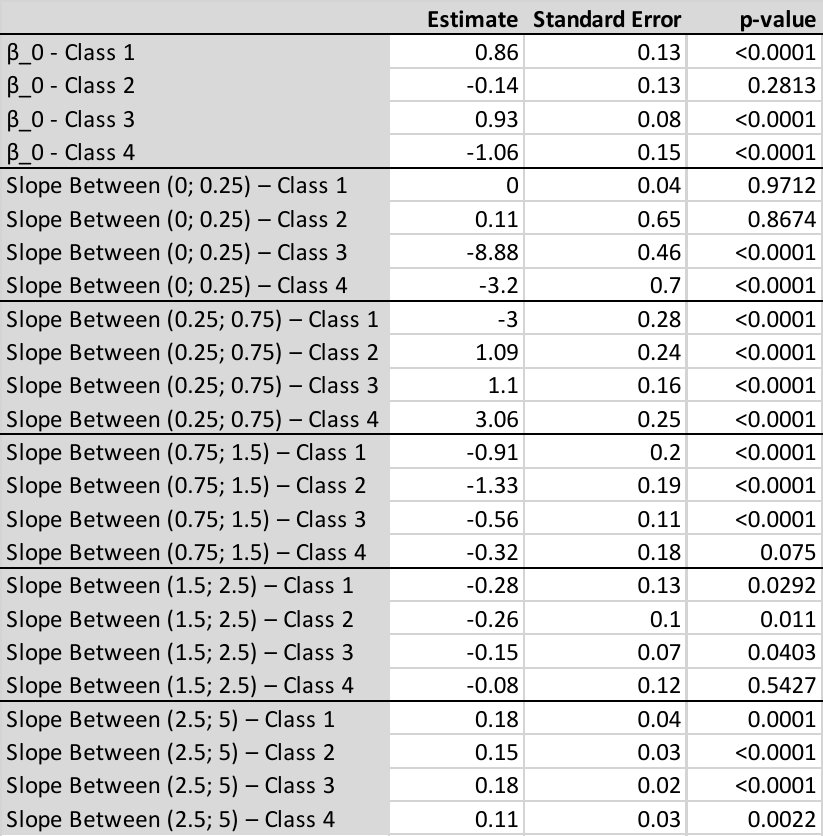

Table A5: Fixed Effect Estimates from the Measurement model within the LCMM framework given the zWeight response.

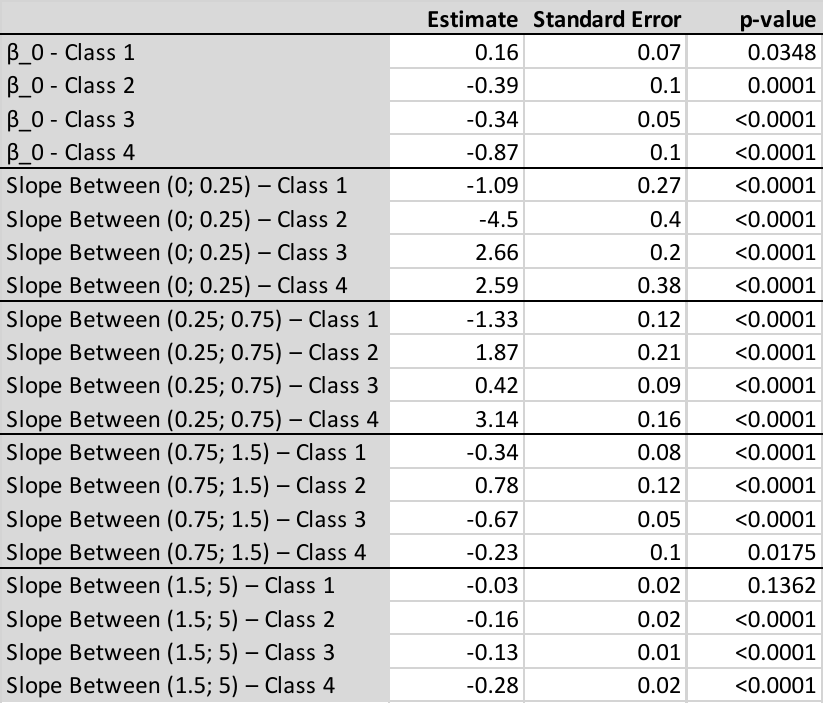

Table A6: Fixed Effect Estimates from the Measurement model within the LCMM framework given the zWFH response.

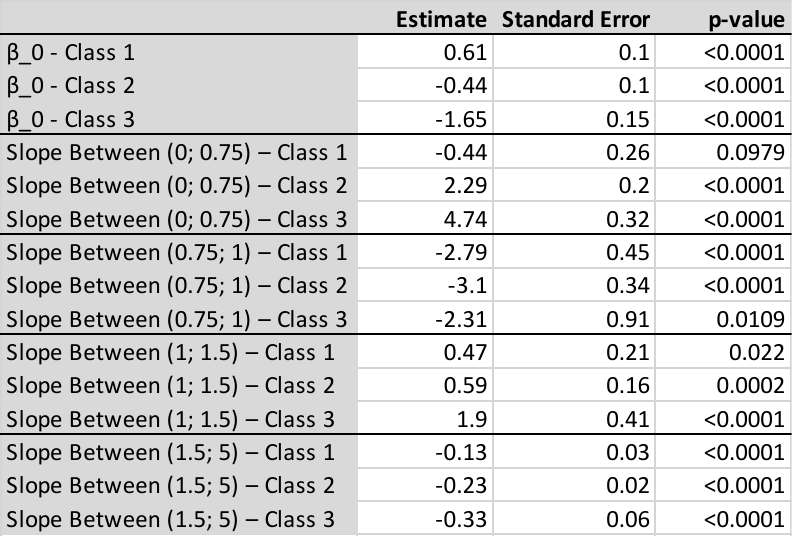

Table A7: Fixed Effect Estimates from the Measurement model within the LCMM framework given the zBMI response.

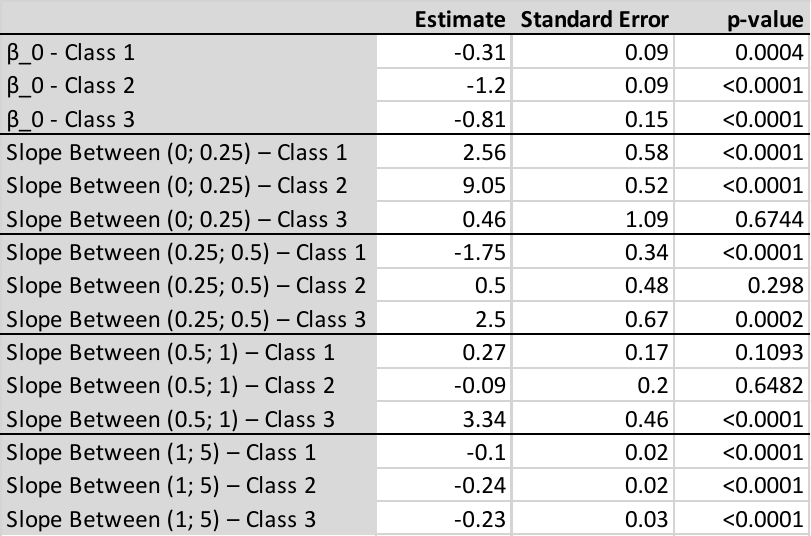

Table A8: Fixed Effect Estimates from the Measurement model within the LCMM framework given the zHeight response within the zHeight+zWeight model.

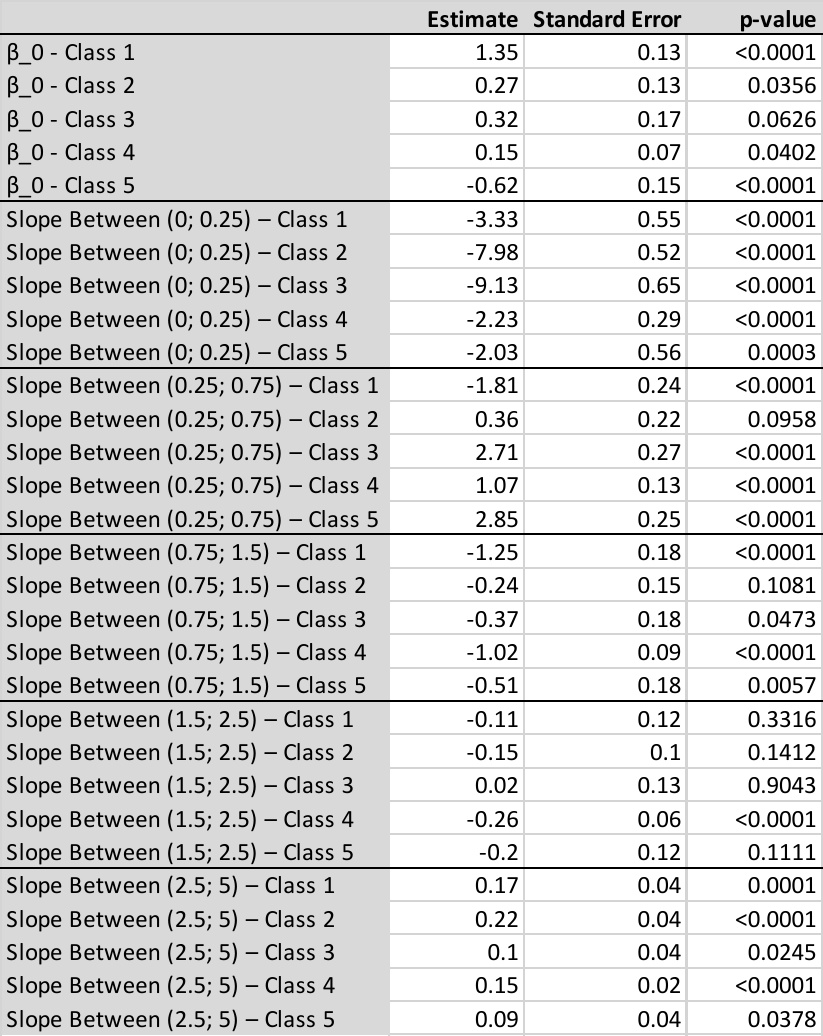

Table A9: Fixed Effect Estimates from the Measurement model within the LCMM framework given the zWeight response within the zHeight+zWeight model.

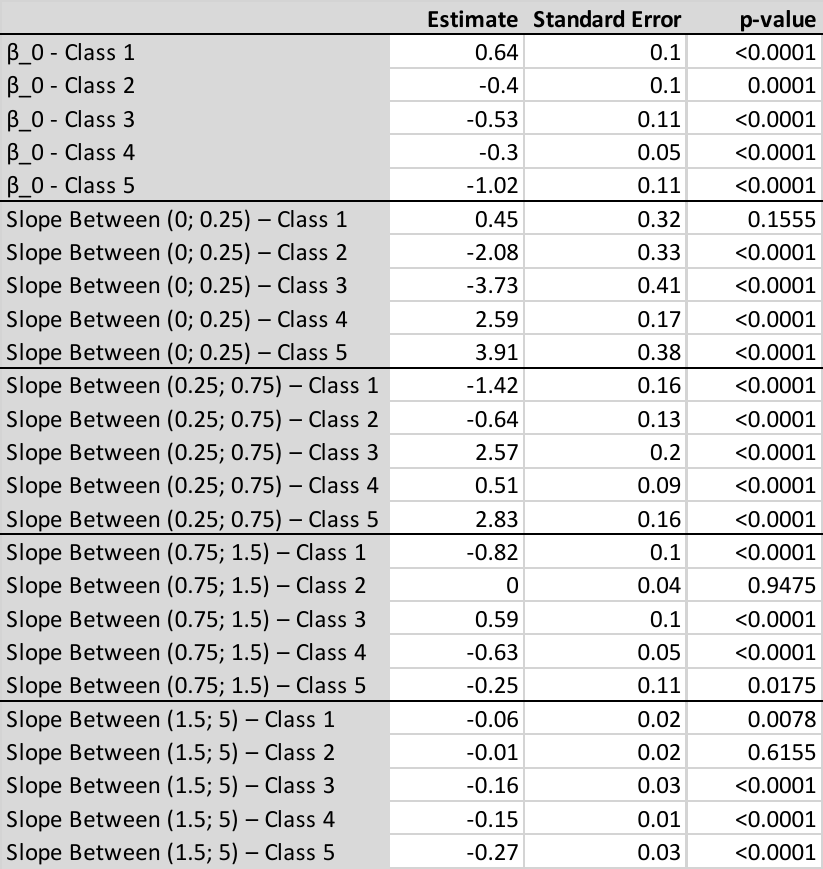

*3.6: Profiles given Increased Knot Locations*

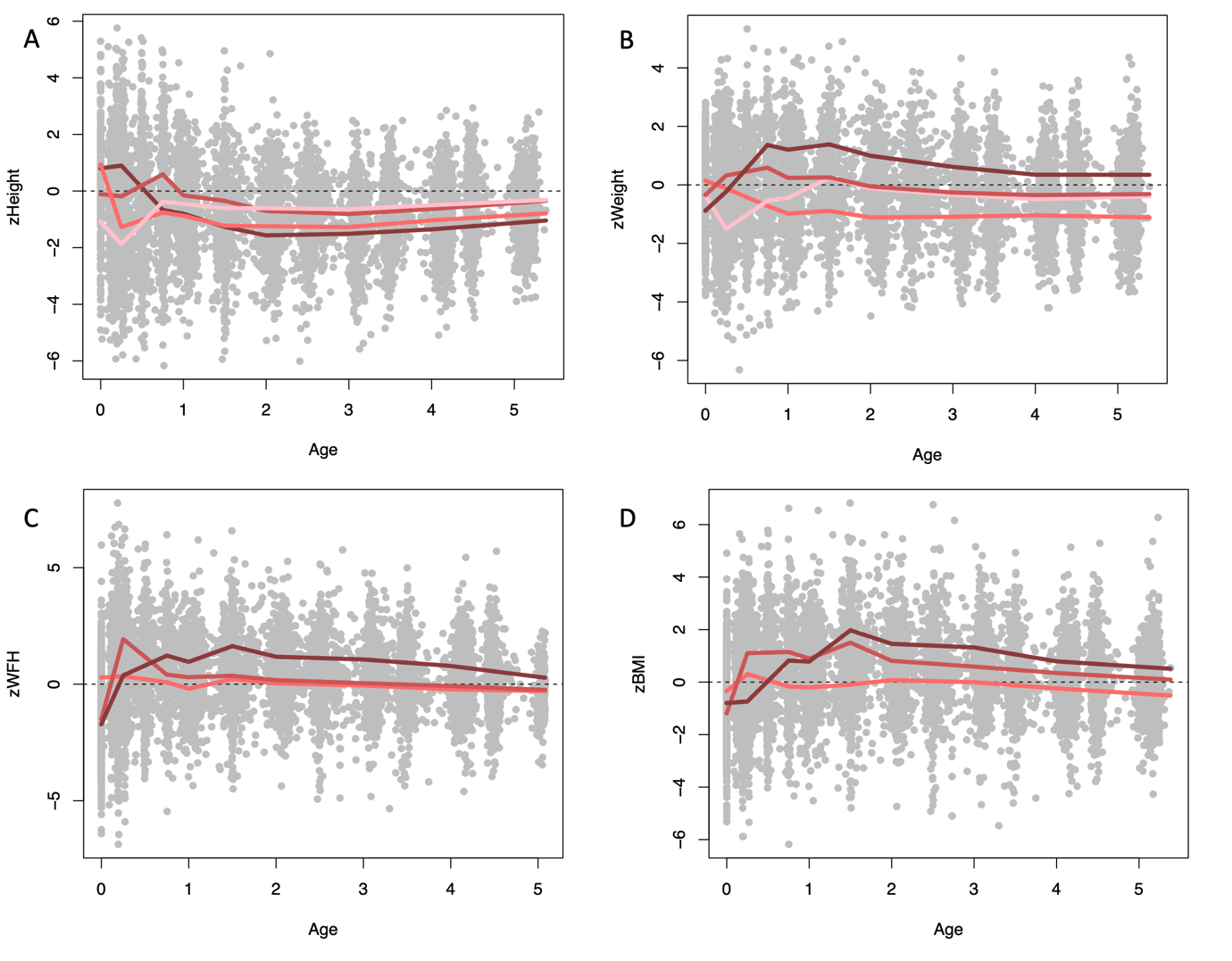
Figure A16: Latent Growth Trajectories identified within A) zHeight, B) zWeight, C) zWFH and D) BMI given additional knots placed at timepoints (0.25,0.75,1,1.5,2,3,4).

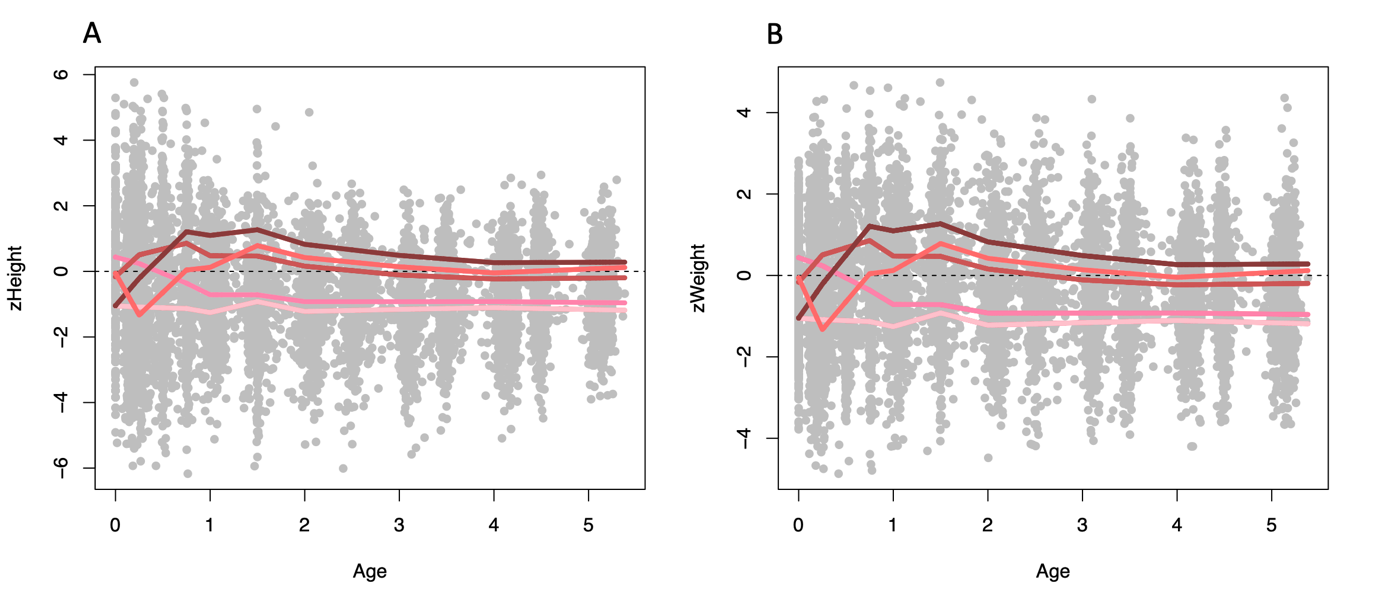
Figure A17: Latent Growth Trajectories identified within A) zHeight and B) zWeight as identified using the multivariate response of zHeight+zWeight, given additional knots placed at timepoints (0.25,0.75,1,1.5,2,3,4).

*3.7: Link Function*

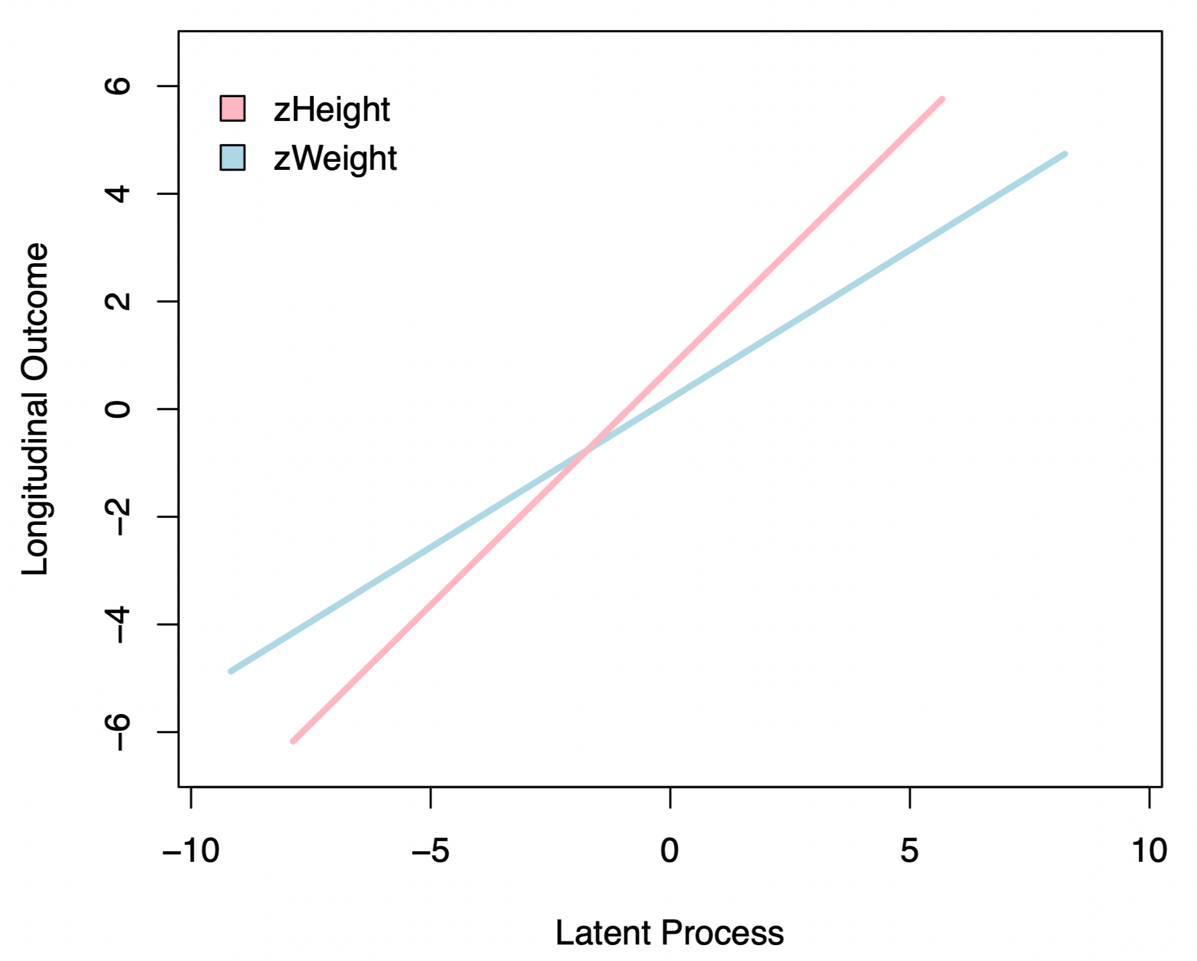

Figure A18: Linear link functions illustrating the relationship between zHeight and zWeight and the Latent Process within the multivariate zHeight+zWeight model.

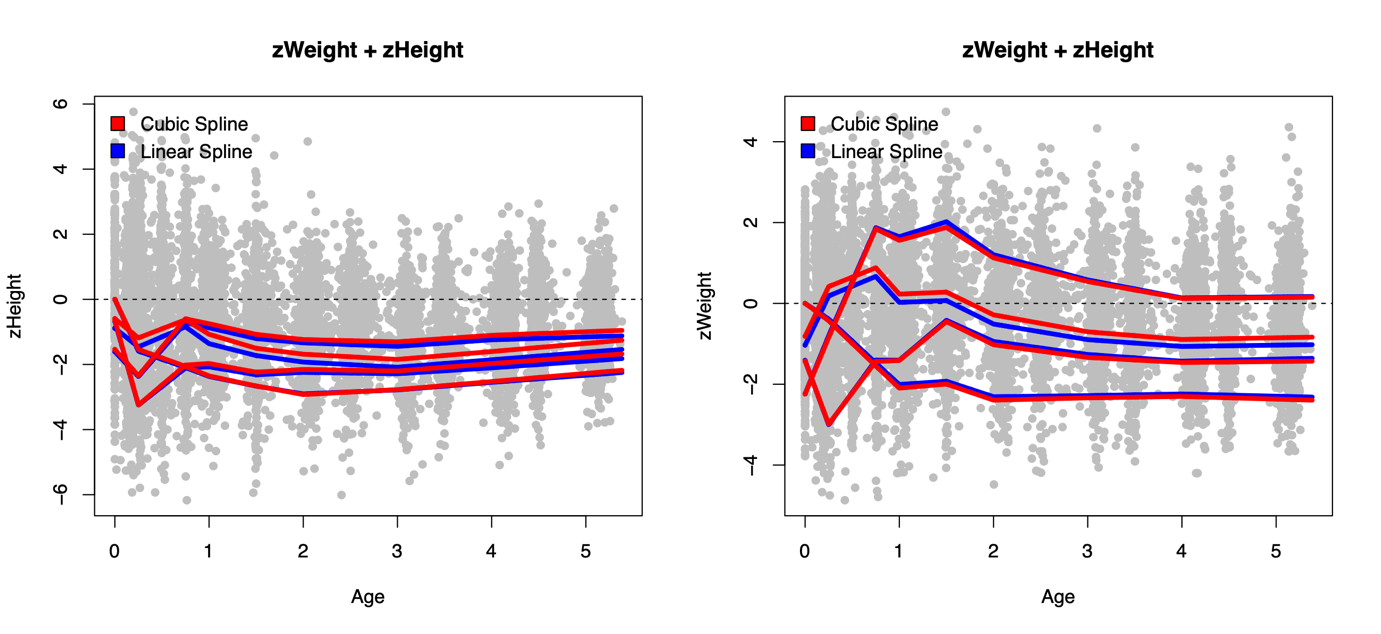

Figure A19: Trajectories of zHeight and zWeight profiles identified given four latent classes when considering a linear or cubic (with three equally spaced knots) spline link function specifying the relationship between the longitudinal outcomes and latent process. Here the effect of link is shown on k=4; when k=5 is considered the cubic spline approach identifies a class of outliers (n=33) which does not meet the criteria for adequate class size, thus leading to k=4 as the optimal number of classes considered.

*3.8: Comparing zBMI and zWFH*

Table A10: Comparison of subject allocations to zBMI and zWFH classes. Comparing these allocations when considering trajectories from birth until 5 years of age.

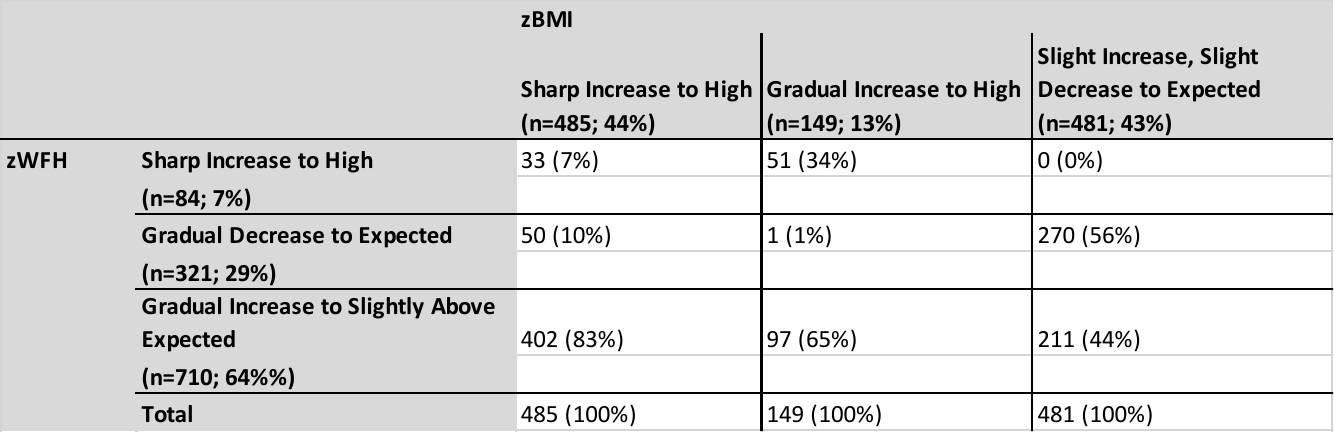

*C9: Rapid Weight Gain*

Table A11: Proportion of children that experienced rapid weight gain within the first 9 months of life within the three zBMI growth classes.

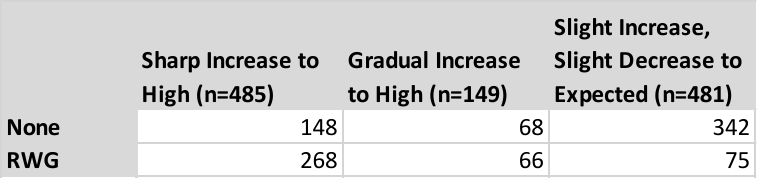

Table A12: Proportion of children that experienced rapid weight gain within the first 9 months of life within the three zWFH growth classes.

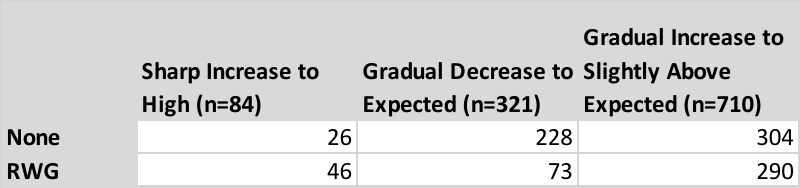

Table A13: Proportion of children that experienced rapid weight gain within the first 9 months of life within the five zHeight+zWeight growth classes.

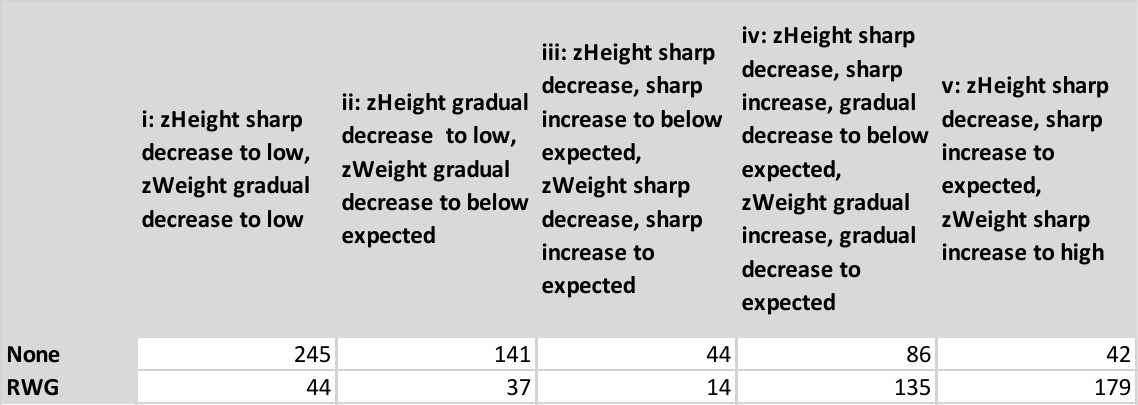
